# Quantifying Normal Diaphragmatic Motion and Shape and their Developmental Changes via Dynamic MRI

**DOI:** 10.1101/2024.05.12.24306850

**Authors:** You Hao, Jayaram K. Udupa, Yubing Tong, Caiyun Wu, Joseph M. McDonough, Samantha Gogel, Oscar H. Mayer, Mostafa Alnoury, Patrick J. Cahill, Jason B. Anari, Drew A. Torigian

## Abstract

**Background:** The diaphragm is a critical structure in respiratory function, yet in-vivo quantitative description of its motion available in the literature is limited.

**Research Question:** How to quantitatively describe regional hemi-diaphragmatic motion and curvature via free-breathing dynamic magnetic resonance imaging (dMRI)?

**Study Design and Methods:** In this prospective cohort study we gathered dMRI images of 177 normal children and segmented hemi-diaphragm domes in end-inspiration and end-expiration phases of the constructed 4D image. We selected 25 points uniformly located on each 3D hemi-diaphragm surface. Based on the motion and local shape of hemi-diaphragm at these points, we computed the velocities and sagittal and coronal curvatures in 13 regions on each hemi-diaphragm surface and analyzed the change in these properties with age and gender.

**Results:** Our cohort consisted of 94 Females, 6-20 years (12.09±3.73), and 83 Males, 6-20 years (11.88±3.57). We observed velocity range: ∼2mm/s to ∼13mm/s; Curvature range –Sagittal: ∼3m^-1^ to ∼27m^-1^; Coronal: ∼6m^-1^ to ∼20m^-1^. There was no significant difference in velocity between genders, although the pattern of change in velocity with age was different for the two groups. Strong correlations in velocity were observed between homologous regions of right and left hemi-diaphragms. There was no significant difference in curvatures between genders or change in curvatures with age.

**Interpretation:** Regional motion/curvature of the 3D diaphragmatic surface can be estimated using free-breathing dynamic MRI. Our analysis sheds light on here-to-fore unknown matters such as how the pediatric 3D hemi-diaphragm motion/shape varies regionally, between right and left hemi-diaphragms, between genders, and with age.

---

Breathing-related motion analysis is important in the study of many disease processes [1]. The diaphragm is the main respiratory muscle involved during inspiration [2, 3] and accounts for ∼70% of the inspired air volume during regular breathing [4]. Thus, the quantification of 3D motion of the diaphragmatic surface is important in a variety of disorders. For example, in patients with thoracic insufficiency syndrome (TIS) [5, 6] and in various pediatric and adult conditions involving scoliosis, diaphragm motion may be altered due to spinal curvature and rotation and chest wall deformities. Equally important but fundamentally lacking is the study and understanding of <u>3D</u> diaphragmatic motion and shape and their developmental changes in <u>normal growing</u> children. Such studies are needed to understand the influence of disease conditions on diaphragm function, to devise appropriate treatment strategies, and to evaluate the effects of therapeutic interventions.

Although studies have been reported in the literature to analyze diaphragmatic motion in a limited manner in different disease conditions based on various imaging techniques [3, 4, 7, 8], to the best of our knowledge, no techniques currently exist for quantifying the full 3D motion and/or 3D shape of the diaphragm in vivo under natural tidal breathing conditions. Therefore, a normative database of the 3D motion and shape of the diaphragm does not exist. In this paper, we present such a method by employing free-breathing dynamic magnetic resonance imaging (dMRI). Our motivation for this approach is that breathing restrictions imposed by imaging requirements, such as breath-holding at different lung volumes, etc., do not reflect the natural state of breathing specific to each subject, and therefore would impede the measurement of diaphragm function and shape during breathing exhibited by the patient during most of their natural living condition.

Compared to computed tomography (CT) and fluoroscopy, dMRI has several advantages, including better soft tissue contrast, lack of ionizing radiation, and flexibility in selecting scanning slice planes. However, limited by current hardware, software, and acquisition techniques, it is difficult to achieve simultaneously high spatial, contrast, and temporal resolution and high image quality for 3D volumetric dMRI. Therefore, we used an approach with fast 2D sequences to first obtain a collection of 2D dMRI images under free-breathing conditions, and then constructed the 4D image representing the thorax over one respiratory cycle for subsequent analysis employing an established technique [9, 10]. In this paper, based on such dMRI data acquired from normal pediatric subjects, we show how motion and shape can be quantified in true 3D/4D space for each hemi-diaphragm to establish, for the first time, in vivo normal 3D diaphragmatic motion and shape measurements during tidal breathing and their changes with child maturation.

## Study Design and Methods

For full details on Methods, please see the supplement document.

### Subject cohort

We performed free-breathing thoracic dMRI scans of 177 healthy children of ages 6 to 20 years with the demographics as in Table 1. Each dMRI study takes ∼40 min per subject and produces ∼2,500 sagittal slices across the chest which constitute a spatiotemporal scanning of the dynamic chest. From this set, a subset of ∼250 slices is selected optimally to represent a 4D image constituting the dynamic thorax over one respiratory cycle, through a previously established 4D image construction method [9]. Subsequently, several image processing operations are applied to the 4D image to derive motion and shape parameters of the diaphragm as outlined below.

**Table 1.**
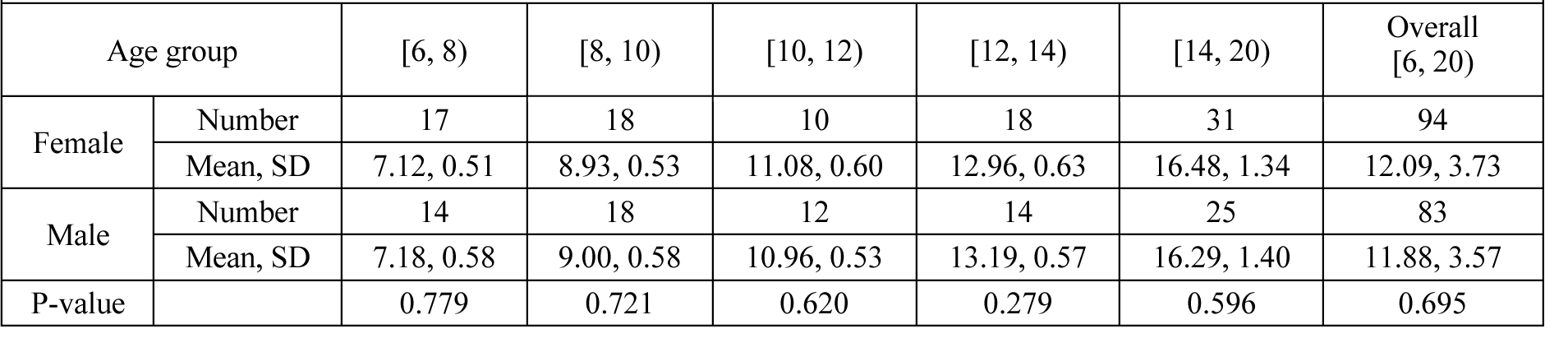
Demographics of our cohort of 177 subjects. Age ranges are indicated by half open intervals. For example, [6, 8) = 6 ≤ age < 8.

| Table 1. Demographics of our cohort of 177 subjects. Age ranges are indicated by half open intervals. For example, [6, 8) = $6 \leq \text{age} < 8$. | | | | | | | |
| --- | --- | --- | --- | --- | --- | --- | --- |
| Age group |  | [6, 8) | [8, 10) | [10, 12) | [12, 14) | [14, 20) | Overall [6, 20) |
| Female | Number | 17 | 18 | 10 | 18 | 31 | 94 |
|  | Mean, SD | 7.12, 0.51 | 8.93, 0.53 | 11.08, 0.60 | 12.96, 0.63 | 16.48, 1.34 | 12.09, 3.73 |
| Male | Number | 14 | 18 | 12 | 14 | 25 | 83 |
|  | Mean, SD | 7.18, 0.58 | 9.00, 0.58 | 10.96, 0.53 | 13.19, 0.57 | 16.29, 1.40 | 11.88, 3.57 |
| P-value |  | 0.779 | 0.721 | 0.620 | 0.279 | 0.596 | 0.695 |

### Diaphragm delineation

Delineation of the diaphragm dome was performed manually in the sagittal slices of the 4D image by a trained operator (Figure 1). A 3D surface of the diaphragm for each respiratory phase was then constructed by using the CAVASS software [11]. Subsequently, the diaphragm was divided manually into right hemi-diaphragm (RHD) and left hemi-diaphragm (LHD) (Figure 2(a)).

**Figure 1.**
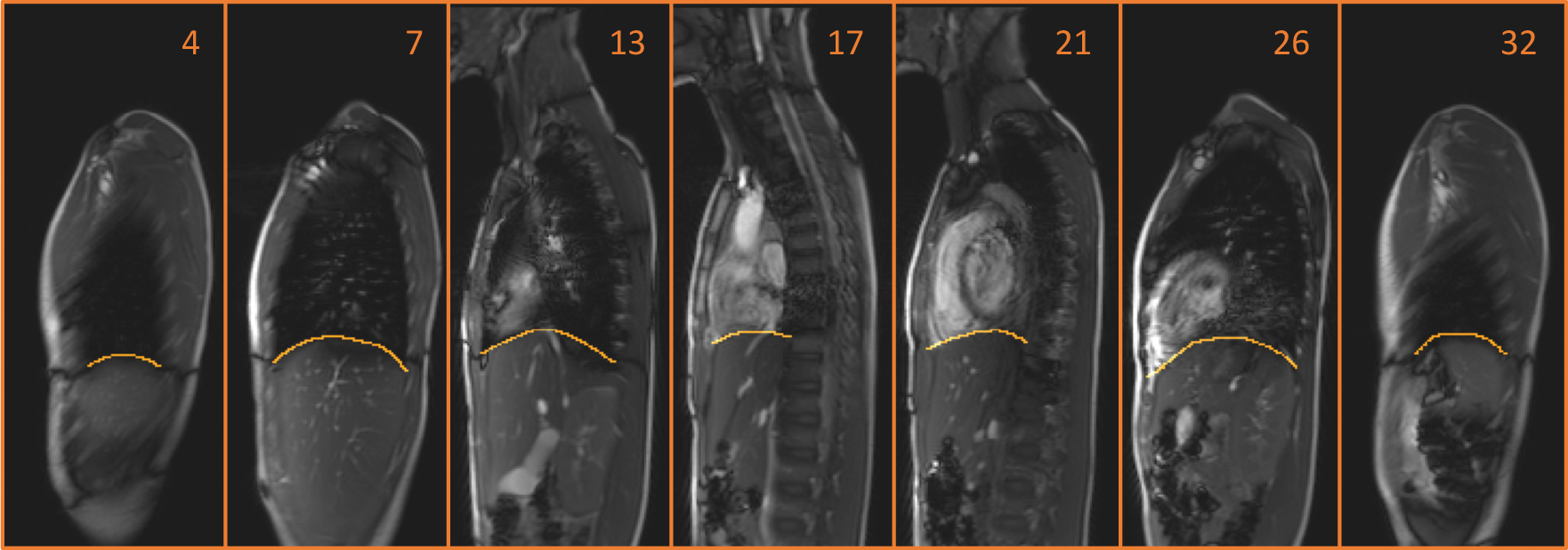
Delineation of the diaphragm in the sagittal plane on the 4D constructed dynamic magnetic resonance images. The displayed number indicates the location of the sagittal slice from right to left at the end-expiration respiratory phase.

**Figure 2.**
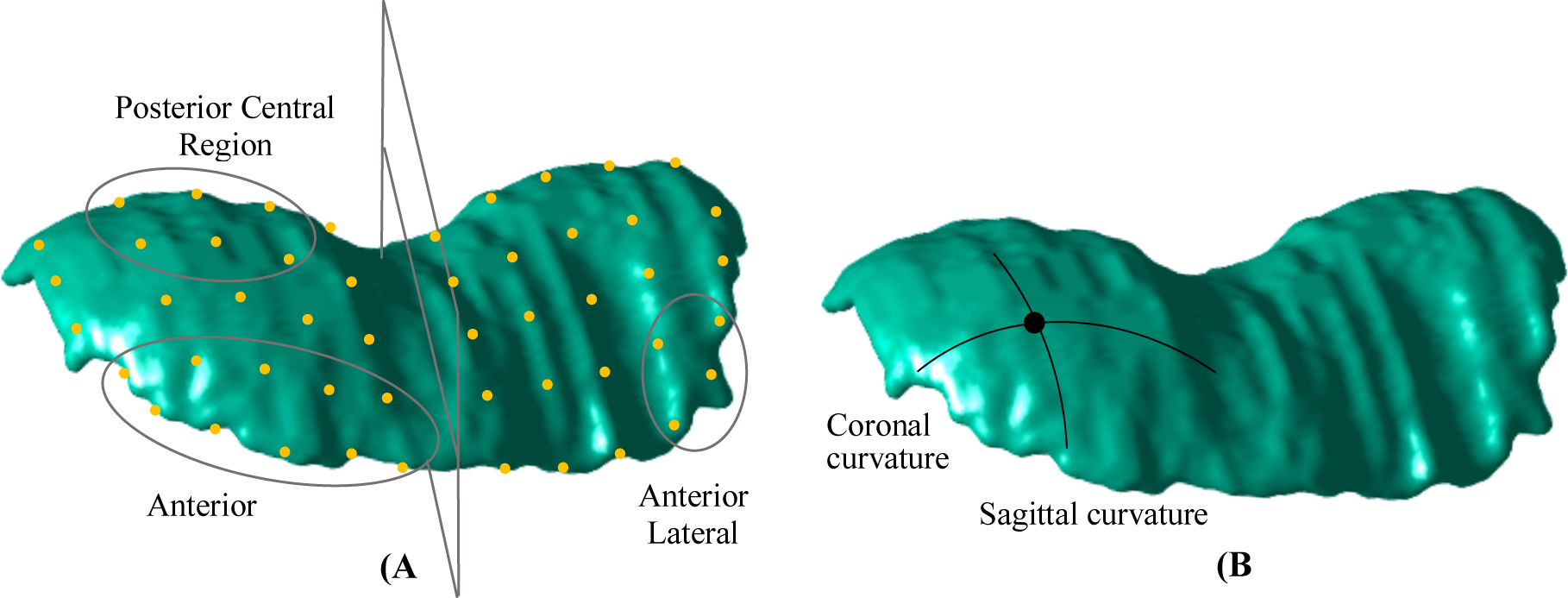
**(A)** The diaphragm surface is divided into left hemi-diaphragm (LHD) and right hemi-diaphragm (RHD) components using the mid-sagittal plane. On each hemi-diaphragm surface, 25 roughly equally-distributed points are selected at each of end-inspiration (EI) and end-expiration (EE) time points. The figure illustrates the EE time point. For describing regional motion and curvature, several regions are defined as collections of points on each hemi-surface. Three exemplar regions are shown – Anterior region and Posterior Central region on the RHD surface and Anterior Lateral region on the LHD surface. **(B)** At each point on each hemi-surface, two types of curvatures are defined – sagittal curvature and coronal curvature. Figure 3. Exemplar plots of velocity vs. age for Posterior Central Region (PCR) of right hemi-diaphragm (RHD) and Anterior Central Region (ACR) of left hemi-diaphragm (LHD). Mean ± one standard deviation limits are indicated.

### Selection of points

To analyze the regional movement of each hemi-diaphragm surface as it excurses from end-expiration (EE) to end-inspiration (EI), we selected 25 points uniformly distributed on each of RHD and LHD at each of EE and EI time points (Figure 2(a)).

### Estimating diaphragm velocities and curvatures

The caudo-cranial displacements of the 25 points from EE to EI were then quantified and the velocity of the RHD and LHD surfaces (in units of mm/s) at each point was derived by dividing the displacement from EE to EI by the time interval from EE to EI.

We describe curvature of the RHD/LHD surface at any point P by two parameters: curvature in the sagittal plane and curvature in the coronal plane (Figure 2(b)). The curvature at P in any plane is simply the reciprocal of the radius of the circle that best approximates the segmented hemi-diaphragm in that plane in the vicinity of P. We express curvature in units of inverse meter (m^-1^).

For facilitating the description of diaphragm motion and curvature regionally, we defined 13 regions on each hemi-surface, where each region is expressed by a subset of the 25 points (Figure 2). In each such region, we expressed an average motion, average sagittal curvature, and average coronal curvature over all points in the region. The definition of these 13 regions can be found in the supplement materials.

### Statistical analysis

We analyzed motion in each of the 13 regions as functions of age and studied how they varied between RHD and LHD surfaces for each gender, as well as how they differed between genders.

Along similar lines, we analyzed sagittal and coronal curvatures in each of the 13 regions at EE and EI separately as functions of age and gender.

## Results

### Hemi-diaphragm velocity

Table 2 summarizes mean and standard deviation of velocity values in the 13 regions of RHD and LHD over the subject cohort for each gender. The P values in the last column indicate the significance level for an unpaired t-test comparison between the entire female group and male group for each region in the RHD and LHD. The other two P-value columns indicate the significance level for a paired t-test comparison between RHD and LHD for each region considering velocities over all subjects for each gender separately. Since the posterior central region (PCR) of the RHD demonstrated the highest velocity over the entire diaphragm (see Table 2), we sought to compare velocities among age groups based on this region’s velocity, as illustrated in Table 3. Supplementary e-Table 1 lists velocities for all regions, age groups and genders. P values less than 0.05 are highlighted.

**Table 2.** Velocity (mm/s) of RHD and LHD measured in 13 regions in our cohort for each gender. Mean (1^st^ value) and standard deviation (2^nd^ value) are listed.

|  |  |  |  |  |  |  |  |  |
| --- | --- | --- | --- | --- | --- | --- | --- | --- |
| Region | Female |  |  | Male |  |  | P value Female vs. Male |  |
|  | RHD | LHD | P value | RHD | LHD | P value | RHD | LHD |
| AR | 5.29, 2.17 | 3.72, 2.98 | **<0.001** | 4.90, 2.16 | 2.68, 1.78 | **<0.001** | 0.236 | **0.006** |
| PR | 10.32, 2.91 | 8.83, 3.21 | **<0.001** | 9.89, 3.30 | 8.10, 2.45 | **<0.001** | 0.355 | 0.096 |
| LR | 7.89, 3.06 | 6.25, 3.25 | **<0.001** | 7.57, 3.22 | 5.89, 2.20 | **<0.001** | 0.499 | 0.400 |
| MR | 6.98, 3.03 | 5.99, 2.87 | **<0.001** | 6.65, 2.74 | 4.82, 2.17 | **<0.001** | 0.452 | **0.003** |
| CR | 8.37, 2.67 | 6.55, 3.05 | **<0.001** | 8.27, 2.62 | 6.01, 2.28 | **<0.001** | 0.796 | 0.195 |
| ALR | 5.84, 2.88 | 4.10, 3.54 | **<0.001** | 5.59, 2.85 | 3.59, 2.28 | **<0.001** | 0.569 | 0.266 |
| AMR | 4.61, 3.21 | 3.48, 3.05 | **<0.001** | 4.21, 3.06 | 2.16, 2.58 | **<0.001** | 0.403 | **0.002** |
| ACR | 5.43, 2.34 | 3.35, 3.34 | **<0.001** | 5.24, 2.49 | 2.42, 2.43 | **<0.001** | 0.591 | **0.037** |
| PLR | 9.80, 3.63 | 8.19, 3.50 | **<0.001** | 9.43, 4.13 | 7.91, 2.76 | **0.002** | 0.517 | 0.557 |
| PMR | 9.74, 3.89 | 8.73, 4.13 | **0.012** | 9.37, 3.56 | 7.62, 3.03 | **<0.001** | 0.511 | **0.044** |
| PCR | 11.48, 3.55 | 9.61, 3.65 | **<0.001** | 11.24, 3.89 | 9.15, 3.37 | **<0.001** | 0.665 | 0.383 |
| CLR | 8.05, 3.25 | 6.50, 3.45 | **<0.001** | 7.72, 3.09 | 6.20, 2.37 | **<0.001** | 0.492 | 0.497 |
| CMR | 6.61, 3.00 | 5.69, 2.77 | **0.002** | 6.41, 2.69 | 4.73, 2.49 | **<0.001** | 0.640 | **0.017** |

RHD = Right Hemi-Diaphragm; LHD=Left Hemi-Diaphragm.

**Table 3.** Mean and standard deviation (SD) of the velocity (mm/s) in the posterior central region (PCR) of the right hemi-diaphragm for different age groups in our cohort. P values indicating the level of significance in a t-test comparison of mean velocities between groups are also listed. Age ranges are indicated by half open intervals. For example, [6, 8) = 6 < age < 8.

| Table 3. Mean and standard deviation (SD) of the velocity (mm/s) in the posterior central region (PCR) of the right hemi-diaphragm for different age groups in our cohort.<br>P values indicating the level of significance in a t-test comparison of mean velocities between groups are also listed. Age ranges are indicated by half open intervals. For example, [6, 8) = 6 ≤ age < 8. |  |  |  |  |  |  |  |  |  |  |
| --- | --- | --- | --- | --- | --- | --- | --- | --- | --- | --- |
|  | Female |  |  |  |  | Male |  |  |  |  |
|  | [6, 8) | [8, 10) | [10, 12) | [12, 14) | [14, 20) | [6, 8) | [8, 10) | [10, 12) | [12, 14) | [14, 20) |
| Mean | 9.22, | 10.35, | 11.24, | 11.32, | 12.69, | 9.16, | 10.74, | 11.62, | 12.98, | 11.33, |
| SD | 1.51 | 3.28 | 1.47 | 2.96 | 3.51 | 1.72 | 2.52 | 4.30 | 3.96 | 4.32 |
| [6, 8) | - | 0.220 | **0.003** | **0.016** | **<0.001** | - | 0.063 | 0.069 | **0.004** | 0.091 |
| [8, 10) | 0.220 | - | 0.429 | 0.364 | **0.029** | 0.063 | - | 0.494 | 0.069 | 0.614 |
| [10, 12) | **0.003** | 0.429 | - | 0.934 | 0.213 | 0.069 | 0.494 | - | 0.418 | 0.852 |
| [12, 14) | **0.016** | 0.364 | 0.934 | - | 0.171 | **0.004** | 0.069 | 0.418 | - | 0.260 |
| [14, 20) | **<0.001** | **0.029** | 0.213 | 0.171 | - | 0.091 | 0.614 | 0.852 | 0.260 | - |

The following is a summary of key results shown in the tables: (1) <u>Female vs. Male:</u> The diaphragm velocity ranges for female and male subjects were [2.5, 12.7] mm/s and [1.6, 12.3] mm/s, respectively, with PCR of RHD for both genders showing maximum velocity and AMR (female) and ACR (male) of LHD showing minimum velocities (e-Table 1). Overall, without regards to subject age, no significant difference in velocity was found between female and male subjects for RHD (Table 2), although for LHD some regions showed statistically significant differences. (2) <u>RHD vs. LHD:</u> For every region, the velocity in RHD was statistically significantly greater than that in the homologous region in LHD, with the greatest difference occurring in ACR for females and males (Table 2). (3) <u>Change with age:</u> Most diaphragm regions did not show a significant change in their velocity with age with the following exceptions: PR, PLR, and PCR showed a significant increase for RHD while CR, ALR, ACR, and CMR showed a significant decrease for LHD. Exemplary plots of velocity vs. age are shown in Figure 3. See Supplementary e-Figure 3 for all such plots for both genders and hemi-diaphragms.

**Figure 3:**
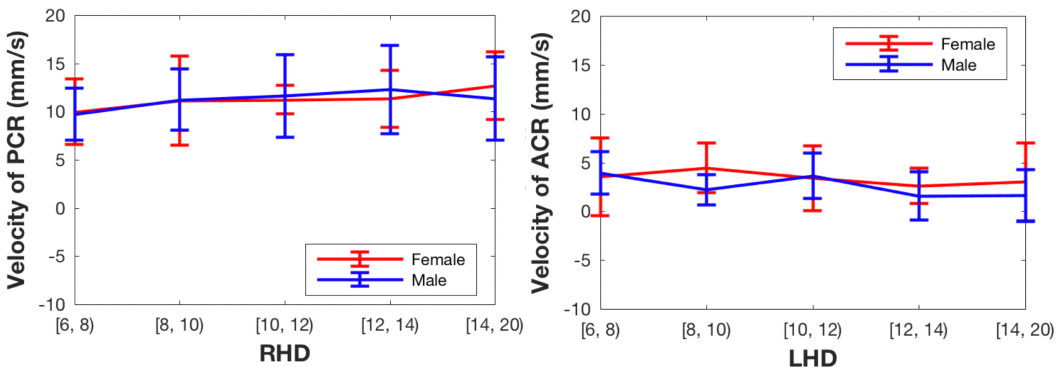
Exemplar plots of velocity vs. age for Posterior Central Region (PCR) of right hemi-diaphragm (RHD) and Anterior Central Region (ACR) of left hemi-diaphragm (LHD). Mean ± one standard deviation limits are indicated.

### Hemi-diaphragm curvatures

Sagittal and coronal curvatures at EE and EI in the 13 regions for RHD and LHD are presented in Supplementary e-Tables 2-5. Table 4 summarizes the change in sagittal curvatures in the 13 regions from EE to EI for RHD and LHD for each gender. Analogously, Table 5 summarizes the change in coronal curvatures. For both curvatures, the signed value representing the change is defined as: curvature at EE – curvature at EI. The P values in the last column indicate the significance level for an unpaired t-test comparison between the entire cohort of female and male subjects for each region separately for RHD and LHD. The other two P-value columns indicate the significance level for a paired t-test comparison between RHD and LHD for each region considering all values over all subjects for each gender separately. Supplemental e-Figures 4-9 further illustrate variations in diaphragm sagittal and coronal curvatures at EE and EI and their changes from EE to EI as a function of age.

**Table 4.** Change in sagittal curvature (m^-1^) of RHD and LHD from end-expiration to end-inspiration measured in 13 regions in our cohort for each gender. Mean (1^st^ value) and standard deviation (2^nd^ value) are listed.

|  |  |  |  |  |  |  |  |  |
| --- | --- | --- | --- | --- | --- | --- | --- | --- |
| Region | Female |  |  | Male |  |  | P value<br>Female vs. Male |  |
|  | RHD | LHD | P value | RHD | LHD | P value | RHD | LHD |
| AR | -2.49, 2.11 | -1.79, 2.23 | **0.011** | -2.35, 2.05 | -1.31, 2.44 | **0.002** | 0.657 | 0.175 |
| PR | 0.69, 1.62 | 1.50, 2.00 | **0.001** | 0.78, 2.17 | 0.70, 1.78 | 0.784 | 0.773 | **0.006** |
| LR | -0.92, 2.22 | 0.30, 2.75 | **<0.001** | -0.81, 2.12 | -0.07, 2.67 | **0.043** | 0.719 | 0.368 |
| MR | -0.96, 2.11 | -0.74, 2.03 | 0.236 | -0.77, 1.91 | -0.67, 2.15 | 0.556 | 0.546 | 0.818 |
| CR | -0.45, 1.17 | -0.24, 1.13 | 0.192 | -0.32, 1.10 | -0.15, 1.40 | 0.409 | 0.452 | 0.616 |
| ALR | -2.85, 3.37 | -1.33, 3.65 | **0.001** | -2.54, 3.50 | -1.47, 4.31 | 0.063 | 0.553 | 0.817 |
| AMR | -2.11, 3.27 | -2.01, 2.90 | 0.784 | -1.97, 3.37 | -1.32, 3.14 | 0.129 | 0.770 | 0.131 |
| ACR | -2.32, 2.23 | -1.98, 2.00 | 0.238 | -2.55, 2.15 | -1.47, 2.21 | **0.002** | 0.492 | 0.111 |
| PLR | -0.12, 2.69 | 1.72, 3.81 | **<0.001** | -0.06, 2.67 | 0.46, 3.02 | 0.23 | 0.881 | **0.016** |
| PMR | 1.58, 3.12 | 1.43, 3.32 | 0.693 | 1.79, 3.84 | 0.87, 3.18 | 0.07 | 0.683 | 0.254 |
| PCR | 1.07, 1.84 | 1.22, 2.33 | 0.643 | 1.13, 2.11 | 1.02, 2.17 | 0.72 | 0.853 | 0.556 |
| CLR | -0.66, 2.84 | 0.35, 3.13 | **0.01** | -0.56, 2.88 | 0.25, 3.18 | 0.09 | 0.814 | 0.835 |
| CMR | -1.57, 3.26 | -1.27, 3.23 | 0.315 | -1.30, 2.91 | -1.13, 3.52 | 0.575 | 0.571 | 0.789 |

RHD = Right Hemi-Diaphragm; LHD=Left Hemi-Diaphragm.

**Table 5.** Change in coronal curvature (m^-1^) of RHD and LHD from end-expiration to end-inspiration measured in 13 regions in our cohort for each gender. Mean (1^st^ value) and standard deviation (2^nd^ value) are listed.

| Region | Female |  |  | Male |  |  | P value<br>Female vs. Male |  |
| --- | --- | --- | --- | --- | --- | --- | --- | --- |
|  | RHD | LHD | P value | RHD | LHD | P value | RHD | LHD |
| AR | -0.19, 2.41 | 0.21, 2.40 | 0.231 | 0.22, 2.38 | 0.30, 2.21 | 0.801 | 0.258 | 0.797 |
| PR | 0.23, 3.00 | 0.72, 2.55 | 0.191 | 0.34, 2.52 | 0.07, 3.03 | 0.509 | 0.803 | 0.123 |
| LR | -1.12, 3.65 | 0.56, 2.36 | **<0.001** | -0.20, 3.52 | 0.27, 3.28 | 0.349 | 0.091 | 0.489 |
| MR | 0.30, 2.87 | 0.47, 3.19 | 0.7 | 1.21, 3.56 | 0.18, 3.69 | 0.075 | 0.059 | 0.575 |
| CR | 0.42, 3.74 | 0.50, 3.47 | 0.877 | 1.08, 4.14 | 1.05, 3.56 | 0.949 | 0.261 | 0.297 |
| ALR | -1.03, 3.54 | -0.09, 3.15 | **0.048** | -0.47, 3.55 | -0.31, 3.44 | 0.76 | 0.299 | 0.664 |
| AMR | 0.48, 4.19 | 0.15, 4.01 | 0.579 | 1.85, 5.54 | 0.09, 4.55 | **0.028** | 0.064 | 0.918 |
| ACR | -0.02, 3.81 | 0.45, 3.85 | 0.373 | 0.63, 4.19 | 0.96, 3.60 | 0.58 | 0.280 | 0.367 |
| PLR | -1.13, 4.17 | 0.70, 2.85 | **<0.001** | -0.14, 4.58 | 0.32, 4.23 | 0.454 | 0.132 | 0.484 |
| PMR | 0.34, 3.42 | 0.89, 4.01 | 0.281 | 0.94, 3.42 | 0.21, 4.24 | 0.197 | 0.250 | 0.277 |
| PCR | 0.45, 4.77 | 1.21, 4.24 | 0.215 | 0.57, 4.02 | 0.71, 4.69 | 0.826 | 0.858 | 0.457 |
| CLR | -1.55, 4.78 | 1.15, 3.66 | **<0.001** | -0.05, 4.50 | 1.00, 4.59 | 0.13 | **0.034** | 0.811 |
| CMR | 0.24, 4.07 | 0.69, 4.24 | 0.456 | 1.02, 4.56 | 0.06, 5.33 | 0.208 | 0.229 | 0.387 |

RHD = Right Hemi-Diaphragm; LHD=Left Hemi-Diaphragm.

We summarize the results from these tables and figures as follows. (1) <u>Female vs. Male:</u> The range of diaphragm curvatures observed were as follows (e-Tables 2-5). Sagittal: [3.1, 26.3] m^-1^ for female and [3.0, 27.2] m^-1^ for male; coronal: [-6.0, 18.9] m^-1^ for female and [-4.8, 19.9] m^-1^ for male. Most regions exhibited no gender-wise difference in sagittal or coronal curvature or the change in curvatures from EE to EI (e-Tables 2-5, Tables 4, 5). (2) <u>RHD vs. LHD:</u> High diaphragm curvatures were observed in central regions (CR, CLR, and CMR for sagittal curvature and CR and CLR for coronal curvature) for both RHD and LHD at both EE and EI (e-Tables 2-5). Most of the regions exhibited statistically significantly different sagittal curvatures between RHD and LHD for both genders (e-Tables 2, 3). For coronal curvatures, the significant difference between RHD and LHD occurred less frequently (e-Tables 4, 5). (3) <u>Change with age:</u> Sagittal curvatures exhibited mostly no change with age at EE (e-Figure 4) but showed reduction at EI for several regions, most markedly for ALR for both hemi-diaphragms (e-Figure 5). Coronal curvatures showed no change with age at EE (e-Figure 6) but showed change with age at EI for LR, PLR, PCR, and CLR for RHD (e-Figure 7).

### Correlations between results

Pearson correlations in velocities between RHD and LHD for the different regions are shown in Supplementary e-Table 6 and between velocities and changes in the two curvatures are summarized in e-Tables 7 and 8, respectively. Table 6 lists the overall correlations that exist between velocity and the change in each type of curvature from EE to EI considering the mean values in each of the 13 regions for each subject while disregarding age. Detailed correlations between velocities and the change in each type of curvature for the different regions, age groups, and genders are provided in Supplementary e-Tables 7 and 8.

**Table 6.** Pearson correlations between velocity and change in sagittal and coronal curvatures of RHD and LHD from end-expiration to end-inspiration measured in 13 regions in our subject cohort for each gender.

| Region | Female |  |  |  | Male |  |  |  |
| --- | --- | --- | --- | --- | --- | --- | --- | --- |
|  | Sagittal |  | Coronal |  | Sagittal |  | Coronal |  |
|  | RHD | LHD | RHD | LHD | RHD | LHD | RHD | LHD |
| AR | -0.07 | -0.28 | -0.1 | -0.11 | -0.12 | **-0.43** | 0.03 | -0.09 |
| PR | 0.29 | 0.15 | 0.09 | 0.27 | 0.31 | 0.15 | 0.37 | 0.31 |
| LR | 0.05 | -0.2 | -0.05 | -0.05 | -0.11 | -0.15 | 0.21 | 0.1 |
| MR | -0.02 | -0.15 | 0.15 | 0.19 | 0.06 | 0.01 | 0.11 | 0.2 |
| CR | 0.22 | 0.24 | 0.12 | 0.34 | 0.03 | 0.31 | **0.48** | **0.49** |
| ALR | -0.06 | -0.21 | 0.02 | 0 | 0 | -0.2 | 0.22 | -0.01 |
| AMR | -0.16 | -0.09 | 0.1 | -0.02 | -0.06 | -0.15 | -0.05 | 0.11 |
| ACR | 0.04 | -0.04 | 0.13 | 0.08 | -0.15 | -0.28 | 0.27 | 0.1 |
| PLR | 0.19 | 0.1 | 0.01 | -0.01 | 0.21 | -0.08 | 0.19 | 0.23 |
| PMR | 0.19 | 0.18 | 0.07 | 0.22 | 0.32 | 0.17 | 0.25 | 0.18 |
| PCR | 0.18 | 0.37 | 0.25 | 0.43 | 0.28 | 0.18 | **0.49** | **0.47** |
| CLR | 0.19 | -0.05 | -0.17 | -0.05 | -0.04 | 0.05 | 0.18 | 0.01 |
| CMR | 0.26 | 0.11 | 0.13 | 0.3 | 0.31 | 0.39 | 0.23 | 0.31 |

RHD = Right Hemi-Diaphragm; LHD=Left Hemi-Diaphragm.

The results in the above tables can be summarized as follows. (1) Velocities in many homologous regions showed strong correlations (>0.6) between RHD and LHD (e-Table 6). (2) Some modest correlations (>0.5) were seen between velocity and change in sagittal curvature from EE to EI for LHD for female subjects (e-Table 7). For RHD, these correlations were generally weak. However, for male subjects, modest correlations were observed for both RHD and LHD and more frequently than for female subjects (e-Table 7). (3) The pattern of velocity correlation between RHD and LHD and velocity to curvature correlation varied considerably between the genders and among age groups for the same region.

Video 1 in the included hyperlink ^1^ shows animations of the dynamic lungs and hemi-diaphragms derived from 4 representative children from our subject cohort: a younger female and an older female, and similarly a younger male and an older male.

## Discussion

Detection, characterization, and quantification of diaphragmatic dysfunction is crucial in the investigation of many disorders which affect respiratory function. Toward such a goal, first understanding normal diaphragmatic motion and shape and the interaction between motion and shape under tidal breathing conditions is essential. We have presented a methodology to accomplish this goal and uncovered the following key results: (i) *Motion*: Velocities in different regions within individual hemi-diaphragms differ considerably, with posterior and anterior regions showing maximum and minimum velocities, respectively, right hemi-diaphragm exhibiting higher velocities than the left, and no significant change in velocity with age. (ii) *Shape*: Curvatures also differ in different regions within individual hemi-diaphragms and between the hemi-diaphragms, with the central regions exhibiting higher curvatures than the peripheral regions. (iii) *Relationship between motion and shape*: High correlations exist for several regions, but not all, between RHD and LHD velocities, and similarly modest correlations exist between velocity and change in curvature from EE to EI. To our knowledge, such a methodology to comprehensively assess diaphragmatic function and structure, and the new information derived from it currently do not exist in the literature.

To illustrate the potential uses of this technology, we present a case study from the TIS application. In Figure 4, we display the lungs and hemi-diaphragm surfaces of a TIS patient pre-operatively (male), with a thoracolumbar congenital scoliosis with the apex centered at the T12/L1 disc, a level of diaphragmatic insertion to the spine. Bilateral rib to pelvis VEPTR (vertical expandable prosthetic titanium rib) devices were placed from T4 and T5 to the pelvis resulting in significant correction of the spinal deformity. The rotated and angulated apex of T12 was now nearly horizontal and translated to midline resulting in a centering of the residual deformity and a close approximation of normal anatomy restoration. The same surfaces of the patient post-operatively, as well as those of normal male subjects, closely matching the pre-and post-operative patient, respectively, are also displayed in Figure 4. The animation of the lungs and hemi-diaphragms for these subjects are available at Video 2 in the provided link^1^ . From a detailed analysis of the mean velocities in the 13 regions of each hemi-diaphragm of these subjects (see e-Table 9), we found that post-operatively, the velocities of all regions increased significantly and overall became closer to those in corresponding regions of matched normal subjects compared to those pre-operatively.

**Figure 4.**
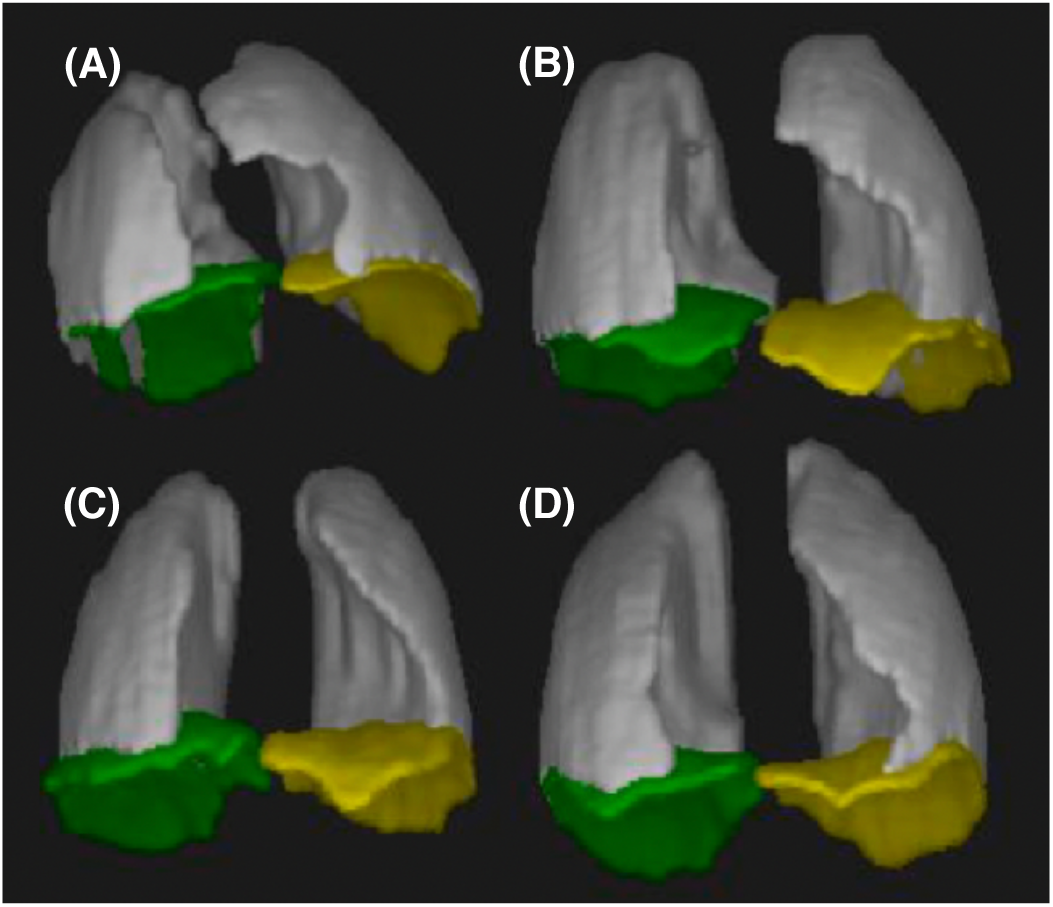
Surface-rendered images of lungs (gray), right hemi-diaphragms (green), and left hemi-diaphragms (yellow) derived from dynamic MRI studies in a pediatric male patient with Thoracic Insufficiency Syndrome (TIS) before **(A)** and after **(B)** surgery, as well as in age-matched male control subjects **(C, D)**.

We may now start seeking answers to key clinical questions like: How do spinal and diaphragmatic deformities restrict individual hemi-diaphragmatic motion and shape, and how are they altered by treatment? Does the percent of deformity correction in EOS or AIS result in normalization of thoracic respiratory function via restoration of diaphragmatic contour?

Several methods for diaphragm measurement and motion analysis have been reported previously [12–22]. Our method offers three key advantages over these earlier methods. 1) Unlike 2D methods [12, 13, 19, 20, 22] which focus only at a couple of 2D planar locations, our approach affords full analysis on the 3D/4D surface of the diaphragm dome and its different local 3D regions. Although some methods acquire full 3D images [14, 18] with excellent temporal resolution, their spatial resolution is poorer than that offered by our approach. Their dynamic acquisitions require breathing maneuvers, and have been demonstrated in adults only and not in children. Children pose special image acquisition and analysis challenges due to their higher respiratory rate and diminished compliance to specific breathing instructions. 2) None of these other methods performed direct motion analysis of points sampled on the 3D surface of the diaphragm but derived indirect indicators of motion. For example, in [12], the vertical distance from the lung apex to the highest point of the diaphragm dome is quantified. In [13], the differential area between the diaphragm curve at different times in the 2D plane is reported. Others have used height, excursion, and antero-posterior size of the diaphragm [15] and volume change [14, 21, 23]. Harlaar et al. [21] evaluated changes in diaphragmatic function in Pompe disease via 3D spoiled-gradient echo (SPGR) breath-hold MRI acquisitions at end-inspiration and end-expiration. However, there was no measurement of point-specific velocity and curvature on the diaphragm dome.

3) None of the existing methods performed diaphragm curvature analysis. Mogalle et al. [18] investigated respiratory muscle weakness through a reduced spatial resolution dynamic 3D MRI series that was controlled with an MR-compatible spirometer. The method does not perform regional motion analysis or curvature estimation and does not demonstrate performance on pediatric subjects.

Our method in its current form has some shortcomings. Although our sample size of 177 is substantial, the number of subjects in younger age groups must be increased, as such subtle differences in group-specific motion and curvature parameters may have been lost in our analysis. We focused only on EE and EI time points in this work, although the method itself is general enough to be applied to any or all time points in the respiratory cycle. Our method relied on manual segmentation of the diaphragm. Finally, we performed detailed analysis only on normal subjects. However, there is no inherent methodological weakness that would prevent us from performing a similar analysis on pediatric patients such as those with TIS as illustrated by the case study in Figure 4.

## Interpretation

We present a unique practical method for performing diaphragm motion and shape analysis based on dynamic MRI acquired under natural breathing conditions. The method allows for motion and curvature analysis on the entire 3D surface of each hemi-diaphragm regionally, investigation of motion and curvature differences among regions and between hemi-diaphragms, and study of the relationship between diaphragmatic motion and shape. Except for manual diaphragm segmentation, all other operations in our method pipeline are automatic and can facilitate easy clinical workflow. Using this methodology, future larger scale prospective studies can be undertaken to quantitatively assess regional and global diaphragmatic dysfunction due to disease conditions and the changes that occur in regional diaphragmatic function and structure due to treatment.

## Data Availability

All data produced in the present study are available upon reasonable request to the authors

All authors declare that they have no conflicts of interest.

## Acknowledgements

**<u>All source(s) of support:</u>** This work is supported by a grant from the National Institutes of Health R01HL150147.

## Acknowledgement

This work is supported by a grant from the National Institutes of Health R01HL150147.

**<u>Take-Home points</u>** <u>(Include 1 sentence for each of the following)</u>

**<u>1) Study Question:</u>** How to quantitatively describe regional hemi-diaphragmatic motion and curvature via free-breathing dynamic magnetic resonance imaging (dMRI)?

**<u>2) Results:</u>** Although there is no significant difference in hemi-diaphragm velocity between genders, the pattern of change in velocity with age is different for the two genders, with strong correlations in velocity between homologous regions of right and left hemi-diaphragms for both genders, and no significant difference in sagittal and coronal curvatures between genders or change in curvatures with age.

**<u>3) Interpretation:</u>** Our analysis sheds light on here-to-fore unknown matters such as how the pediatric normal 3D hemi-diaphragm motion/shape varies regionally, between right and left hemi-diaphragms, between genders, and with age.

## Footnotes

1 http://www.mipg.upenn.edu/Vnews/FileShare/DiaphragmMotion/Videos-Diaphragm-Motion-Shape.pptx

