## Supplemental File for "Quantifying Normal Diaphragmatic Motion and Shape and their Developmental Changes via Dynamic MRI"

**Running title:** Quantification of diaphragmatic motion and shape

### Supplement

#### Details on Methods

##### Subject cohort

We enrolled 177 healthy children of ages 6 to 20 years (with the demographics as in Table 1) with the following exclusion criteria: (i) history of thoracic surgery; (ii) history of asthma or other lung disease; (iii) respiratory tract illness within the last 30 days; and (iv) history of scoliosis or other congenital skeletal abnormality. We performed thoracic dMRI scans of these children with the following scan protocol: 3T MRI scanner (Siemens Healthcare, Erlangen, Germany), true-FISP imaging with steady-state precession sequence; TR/TE = 3.82/1.91 msec; voxel size approximately 1 x 1 x 6 $mm^3$; 320 x 320 matrix; bandwidth = 558 Hz; and flip angle = 76°. For each subject, 30-40 2D sagittal slices at contiguous locations across the entire chest were acquired, and for each sagittal location, 80 slices were acquired continuously at a rate of ~480 ms per slice under free-breathing conditions. Each study produced a total of ~2,500 sagittal slices in this manner taking a total scan time of ~40 min. All data were acquired under an ongoing research protocol approved by the Institutional Review Board at the Children's Hospital of Philadelphia (CHOP) and University of Pennsylvania, along with Health Insurance Portability and Accountability Act waiver.

A subset of ~250 slices is selected optimally from the set of ~2,500 scanned slices to represent a 4D image constituting the dynamic thorax over a typical respiratory cycle. The optimal selection is done through a previously established 4D image construction method which has been validated via a dynamic 3D-printed lung phantom [E1, E2].

##### Diaphragm delineation

Delineation of the diaphragm dome (along its superior aspect) was subsequently performed manually in the sagittal slices of the 3D images corresponding to end-expiration (EE) and end-inspiration (EI) time points of the constructed 4D image by a trained operator as illustrated in Figure 1. The diaphragm was delineated from the anterior-most aspect to the posterior-most aspect, including portions adjacent to other organs/tissue structures such as the heart, liver, or mediastinal fat, on all slices passing through the diaphragm. A 3D surface of the diaphragm for each respiratory time point was then constructed by using the CAVASS software [E3]. Subsequently, the diaphragm was divided manually into right hemi-diaphragm (RHD) and left hemi-diaphragm (LHD) by choosing the midline sagittal slice as the dividing plane as illustrated in Figure 2(a).

**Selection of points**

The goal of our method was to analyze the regional movement of each hemi-diaphragm surface from EE to EI. To facilitate this, we selected 25 points uniformly distributed on each of RHD and LHD surfaces at each of EE and EI time points as follows (Figure 2(a)). We first selected 5 evenly spaced sagittal slices in the 3D image corresponding to EE and EI in the medio-lateral direction that passed through each hemi-diaphragm surface. Then, in each of these sagittal slices, we selected 5 evenly-spaced points in the anterior-posterior direction along the delineated diaphragm contour. The caudo-cranial displacements of these 25 points during the breathing cycle from EE to EI were then quantified.

**Estimating diaphragm velocities**

Since the time interval from EE to EI in the 4D constructed image is known for each subject, we obtained the velocity of the RHD and LHD surfaces (in units of mm/s) at each of the 25 points by dividing the displacement from EE to EI by the time interval from EE to EI. Although, for this initial exploration, we utilized only the EE and EI time points, the approach can be employed in the future to estimate diaphragmatic velocity over the entire respiratory cycle at any respiratory phase.

**Estimating diaphragm curvatures**

Curvature at different points on the RHD and LHD surfaces is an important indicator of shape and shape change with respiratory motion. Intuitively, curvature of a 2D curve is the amount by which the curve deviates from a straight line. Formally, the curvature $\kappa$ at a point P on a 2D smooth curve is defined as the reciprocal, $\kappa = 1/R$, of the radius $R$ of the circle that best approximates the curve at point P. The radius $R$ of the best-fitting circle can be obtained by derivatives of the curve evaluated at point P. The larger the radius of the best-fit circle, the lower is the curvature, and the smaller the radius, the higher is the curvature (see e-Figure 1 in the supplement). In order to make the numerical values reasonable, we express curvature in units of inverse meter ($m^{-1}$) in this paper. Otherwise, if expressed in $cm^{-1}$ or $mm^{-1}$, numerically the values will be very small decimal numbers.

When applying the above curvature concept from 2D curves to the three-dimensional RHD and LHD surfaces, first we smooth the diaphragm boundary. This is because the delineated curves are not smooth locally due to their digital nature (e-Figure 2). Secondly, there are different ways of describing the 3D analogue of curvature defined above for 2D planar curves on a 3D surface. Any such description has two components of curvature. Here, we define the two components by considering curvature separately in the sagittal plane and the coronal plane (Figure 2(b)) at point P, since this is anatomically more meaningful and interpretable than other possible definitions such as principal curvatures at P. We first obtain the two curves from the smoothed surface, and then apply the above 2D definition to each curve to compute curvature.

**Describing diaphragm motion and curvature**

For facilitating the description of diaphragm motion and curvature regionally, rather than describing at each of the 50 points on the diaphragmatic surface, we divided each hemi-surface into 13 regions, where each region is expressed by a subset of the 25 points as follows (see Figure 2). In each such region, we expressed an average motion, average sagittal curvature, and average coronal curvature over all points in the region. The 13 regions are defined as follows: 1) *Anterior Region* (AR): 10 points (2 (rows) x 5 (columns)) from the 2 (coronally-oriented) rows through the anterior hemi-diaphragm (see Figure 2(a)); 2) *Posterior Region*

(PR): 10 points (2x5) from the 2 rows through the posterior hemi-diaphragm; 3) *Lateral Region* (LR): 10 points (5x2) from the 2 (sagittally-oriented) columns through the lateral hemi-diaphragm; 4) *Medial Region* (MR): 10 points (5x2) from the 2 columns through the medial hemi-diaphragm; 5) *Central Region* (CR): 9 points (3x3) in the center of the hemi-diaphragm; 6) *Anterior-Lateral Region* (ALR): 4 points (2x2) closest to the anterior and lateral aspects of the hemi-diaphragm (see Figure 2(a)); 7) *Anterior-Medial Region* (AMR): 4 points (2x2) closest to the anterior and medial aspects of the hemi-diaphragm; 8) *Anterior-Central Region* (ACR): 6 points (2x3) in the anterior 2 rows and central 3 columns through the hemi-diaphragm; 9) *Posterior-Lateral Region* (PLR): 4 points (2x2) closest to the posterior and lateral aspects of the hemi-diaphragm; 10) *Posterior-Medial Region* (PMR): 4 points (2x2) closest to the posterior and medial aspects of the hemi-diaphragm; 11) *Posterior-Central Region* (PCR): 6 points (2x3) in the posterior 2 rows and central 3 columns through the hemi-diaphragm (see Figure 2(a)); 12) *Central-Lateral Region* (CLR): 6 points (3x2) in the central 3 rows and the 2 columns through the lateral hemi-diaphragm; 13) *Central-Medial Region* (CMR): 6 points (3x2) in the central 3 rows and the 2 columns through the medial hemi-diaphragm.

**Statistical analysis**

We sought answers to the following key questions: How do the diaphragm velocities differ in different regions, between RHD and LHD, between genders, and with child maturation? We analyzed motion in each of the 13 regions as functions of age and studied how they varied between RHD and LHD surfaces for each gender, as well as how they differed between genders. We categorized subjects in our cohort into 5 age groups as in Table 1.

Along similar lines, we analyzed diaphragm sagittal and coronal curvatures in each of the 13 regions at EE and EI separately as functions of age. We investigated gender- and age-group-specific differences in their magnitude and direction and differences between LHD and RHD. We also performed several types of analysis of correlation for the 13 regions: between motion of RHD and LHD, between motion of LHD and its sagittal and coronal curvatures, and between motion of RHD and its sagittal and coronal curvatures. Some key results are presented below. Further details are included in the supplement document.

<u>**Details on Results**</u>

This section contains e-Tables 1-9 and e-Figures 1-9 referred to in the main body of the paper.

e-Table 1. Velocity (in mm/s) of right hemi-diaphragm and left hemi-diaphragm measured in 13 regions in our cohort for each gender and age group. Age ranges are indicated by half open intervals. For example, [6, 8) = 6 ≤ age < 8. P values less than 0.05 are highlighted. P values in the last column indicate the level of significance of a paired t-test comparison of velocities of each region between RHD and LHD disregarding age.

**Female**

| **Region** | **Right Hemi-Diaphragm** | | | | | **Left Hemi-Diaphragm** | | | | | **P value** |
|---|---|---|---|---|---|---|---|---|---|---|---|
| | [6, 8) | [8, 10) | [10, 12) | [12, 14) | [14, 20) | [6, 8) | [8, 10) | [10, 12) | [12, 14) | [14, 20) | |
| AR | 5.51, 2.28 | 6.10, 3.02 | 5.77, 1.27 | 5.01, 2.11 | 4.68, 1.67 | 3.84, 3.57 | 4.86, 3.18 | 3.79, 2.60 | 3.04, 1.66 | 3.36, 3.19 | **<0.001** |
| PR | 9.23, 3.46 | 10.09, 3.59 | 9.85, 1.02 | 10.39, 3.54 | 11.16, 1.93 | 8.67, 4.13 | 9.31, 4.42 | 8.38, 2.22 | 8.28, 2.62 | 9.08, 2.44 | **<0.001** |
| LR | 6.96, 4.86 | 8.11, 3.05 | 8.41, 2.08 | 8.25, 2.81 | 7.90, 2.17 | 5.80, 3.20 | 7.31, 4.46 | 5.77, 1.72 | 5.82, 2.59 | 6.27, 3.23 | **<0.001** |
| MR | 6.97, 2.83 | 7.64, 4.66 | 6.60, 2.08 | 6.68, 3.24 | 6.87, 2.04 | 6.17, 3.60 | 7.01, 4.16 | 5.71, 2.65 | 5.53, 1.82 | 5.63, 1.97 | **<0.001** |
| CR | 7.78, 2.37 | 8.57, 3.36 | 8.57, 0.94 | 8.27, 2.39 | 8.55, 2.99 | 6.91, 4.25 | 7.10, 3.64 | 6.83, 2.61 | 5.91, 2.15 | 6.29, 2.54 | **<0.001** |
| ALR | 5.12, 4.38 | 6.38, 2.72 | 6.83, 1.93 | 6.19, 2.86 | 5.39, 2.14 | 4.00, 3.08 | 5.70, 3.97 | 3.24, 2.28 | 3.57, 2.85 | 3.80, 4.07 | **<0.001** |
| AMR | 5.33, 2.85 | 6.07, 5.01 | 4.86, 2.03 | 3.70, 2.01 | 3.80, 2.65 | 3.49, 3.07 | 4.96, 3.98 | 4.20, 2.95 | 2.53, 2.37 | 2.91, 2.61 | **<0.001** |
| ACR | 5.65, 1.74 | 6.03, 3.00 | 6.21, 1.23 | 5.16, 2.02 | 4.86, 2.59 | 3.53, 3.98 | 4.48, 2.54 | 3.38, 3.32 | 2.59, 1.81 | 3.02, 4.00 | **<0.001** |
| PLR | 8.57, 5.67 | 9.57, 3.71 | 10.00, 2.39 | 10.21, 3.28 | 10.31, 2.61 | 7.40, 3.44 | 8.64, 5.00 | 7.90, 1.55 | 7.94, 2.58 | 8.59, 3.52 | **<0.001** |
| PMR | 8.89, 3.01 | 9.78, 5.34 | 8.64, 2.63 | 9.92, 5.24 | 10.43, 2.65 | 8.80, 4.73 | 9.79, 5.45 | 7.31, 2.80 | 8.50, 3.67 | 8.66, 3.57 | **0.012** |
| PCR | 9.96, 3.40 | 11.12, 4.60 | 11.23, 1.47 | 11.32, 2.96 | 12.69, 3.51 | 9.90, 4.60 | 9.87, 4.36 | 9.63, 2.82 | 8.66, 3.07 | 9.83, 3.29 | **<0.001** |
| CLR | 7.27, 4.96 | 8.50, 3.11 | 8.35, 2.28 | 8.39, 3.31 | 7.92, 2.42 | 6.08, 3.43 | 7.58, 4.63 | 6.23, 1.57 | 6.10, 3.19 | 6.42, 3.32 | **<0.001** |
| CMR | 6.83, 2.76 | 6.96, 4.26 | 6.34, 2.14 | 6.68, 3.60 | 6.33, 2.16 | 6.15, 3.78 | 6.09, 3.68 | 5.85, 2.35 | 5.62, 1.87 | 5.17, 2.10 | **0.002** |

**Male**

The 3rd value in each cell indicates the level of significance in a t-test comparison of velocities between female and male subjects for each age group.

| Region | Right Hemi-Diaphragm | | | | | Left Hemi-Diaphragm | | | | | P value |
|---|---|---|---|---|---|---|---|---|---|---|---|
| | [6, 8) | [8, 10) | [10, 12) | [12, 14) | [14, 20) | [6, 8) | [8, 10) | [10, 12) | [12, 14) | [14, 20) | |
| AR | 4.67, 1.34<br>0.233 | 5.07, 1.30<br>0.192 | 4.49, 1.01<br>**0.016** | 4.44, 3.24<br>0.556 | 5.33, 2.69<br>0.272 | 3.49, 1.40<br>0.728 | 2.47, 1.25<br>**0.006** | 3.59, 1.82<br>0.834 | 1.98, 2.12<br>0.124 | 2.30, 1.86<br>0.147 | **<0.001** |
| PR | 8.70, 2.33<br>0.633 | 9.85, 2.66<br>0.820 | 9.81, 3.55<br>0.975 | 10.66, 3.54<br>0.835 | 10.18, 3.92<br>0.227 | 7.71, 2.07<br>0.438 | 8.24, 1.87<br>0.348 | 8.21, 2.43<br>0.867 | 8.78, 3.83<br>0.664 | 7.77, 2.14<br>**0.040** | **<0.001** |
| LR | 6.86, 1.55<br>0.944 | 7.56, 2.73<br>0.570 | 6.56, 2.52<br>**0.079** | 7.59, 3.00<br>0.524 | 8.45, 4.41<br>0.541 | 6.12, 1.97<br>0.743 | 5.87, 2.35<br>0.232 | 6.23, 1.92<br>0.565 | 5.73, 2.66<br>0.920 | 5.69, 2.22<br>0.453 | **<0.001** |
| MR | 5.98, 2.00<br>0.279 | 6.90, 2.05<br>0.544 | 7.06, 2.79<br>0.671 | 6.53, 3.77<br>0.905 | 6.69, 2.97<br>0.786 | 4.99, 1.62<br>0.268 | 5.03, 2.03<br>**0.078** | 4.99, 2.44<br>0.511 | 5.13, 2.32<br>0.585 | 4.30, 2.38<br>**0.026** | **<0.001** |
| CR | 7.57, 1.63<br>0.782 | 8.26, 2.09<br>0.747 | 8.83, 2.61<br>0.774 | 8.84, 3.13<br>0.560 | 8.05, 3.11<br>0.537 | 6.55, 1.65<br>0.770 | 6.32, 2.40<br>0.453 | 6.77, 2.20<br>0.952 | 5.83, 2.57<br>0.931 | 5.22, 2.27<br>0.105 | **<0.001** |
| ALR | 5.26, 1.74<br>0.906 | 5.91, 2.30<br>0.583 | 4.39, 1.73<br>**0.005** | 5.26, 2.82<br>0.365 | 6.30, 3.91<br>0.274 | 4.31, 2.33<br>0.761 | 3.38, 2.31<br>**0.040** | 3.85, 2.14<br>0.525 | 3.01, 2.52<br>0.566 | 3.53, 2.24<br>0.772 | **<0.001** |
| AMR | 3.86, 1.83<br>0.107 | 4.38, 1.41<br>0.177 | 4.25, 1.99<br>0.488 | 3.51, 5.12<br>0.888 | 4.65, 3.48<br>0.305 | 2.79, 1.49<br>0.438 | 1.82, 2.75<br>**0.009** | 3.27, 2.89<br>0.466 | 1.85, 3.28<br>0.501 | 1.67, 2.30<br>**0.066** | **<0.001** |
| ACR | 4.91, 1.73<br>0.247 | 5.34, 1.53<br>0.388 | 5.76, 1.73<br>0.501 | 4.70, 3.81<br>0.665 | 5.38, 2.88<br>0.479 | 3.93, 2.19<br>0.737 | 2.20, 1.51<br>**0.002** | 3.65, 2.34<br>0.824 | 1.54, 2.46<br>0.175 | 1.61, 2.64<br>0.136 | **<0.001** |
| PLR | 8.20, 2.45<br>0.824 | 9.10, 3.45<br>0.697 | 8.54, 3.54<br>0.281 | 9.81, 3.87<br>0.757 | 10.54, 5.47<br>0.837 | 7.62, 2.42<br>0.845 | 8.10, 2.73<br>0.692 | 8.34, 2.52<br>0.639 | 8.01, 3.62<br>0.950 | 7.66, 2.72<br>0.282 | **0.002** |
| PMR | 8.24, 3.00<br>0.556 | 9.91, 3.21<br>0.927 | 10.25, 4.63<br>0.341 | 9.59, 3.92<br>0.847 | 9.05, 3.39<br>0.093 | 7.58, 2.56<br>0.392 | 8.01, 2.50<br>0.218 | 6.82, 3.25<br>0.716 | 8.24, 4.15<br>0.853 | 7.36, 2.91<br>0.148 | **<0.001** |
| PCR | 9.72, 2.68<br>0.827 | 11.21, 3.17<br>0.948 | 11.61, 4.30<br>0.792 | 12.29, 4.58<br>0.472 | 11.33, 4.32<br>0.200 | 9.00, 3.07<br>0.536 | 9.09, 2.40<br>0.509 | 9.63, 3.38<br>0.999 | 9.97, 5.31<br>0.388 | 8.56, 2.84<br>0.134 | **<0.001** |
| CLR | 7.22, 1.48<br>0.974 | 7.72, 2.96<br>0.446 | 6.79, 2.79<br>0.171 | 7.92, 2.91<br>0.675 | 8.33, 4.02<br>0.635 | 6.49, 2.02<br>0.698 | 6.18, 2.69<br>0.274 | 6.46, 1.83<br>0.758 | 6.10, 2.65<br>0.997 | 5.96, 2.54<br>0.569 | **<0.001** |
| CMR | 5.89, 1.75<br>0.279 | 6.44, 2.28<br>0.648 | 6.84, 2.54<br>0.622 | 6.74, 3.86<br>0.968 | 6.28, 2.85<br>0.943 | 4.84, 2.02<br>0.254 | 5.34, 1.78<br>0.441 | 5.12, 2.38<br>0.479 | 5.38, 2.99<br>0.787 | 3.65, 2.72<br>**0.021** | **<0.001** |

RHD = Right Hemi-Diaphragm; LHD=Left Hemi-Diaphragm.

1) *Anterior Region* (AR): 10 points (2 (rows) x 5 (columns)) from the 2 (coronally-oriented) rows through the anterior hemi-diaphragm (see Figure 2(a)); 2) *Posterior Region* (PR): 10 points (2x5) from the 2 rows through the posterior hemi-diaphragm; 3) *Lateral Region* (LR): 10 points (5x2) from the 2 (sagittally-oriented) columns through the lateral hemi-diaphragm; 4) *Medial Region* (MR): 10 points (5x2) from the 2 columns through the medial hemi-diaphragm; 5) *Central Region* (CR): 9 points (3x3) in the center of the hemi-diaphragm; 6) *Anterior-Lateral Region* (ALR): 4 points (2x2) closest to the anterior and lateral aspects of the hemi-diaphragm (see Figure 2(a)); 7) *Anterior-Medial Region* (AMR): 4 points (2x2) closest to the anterior and medial aspects of the hemi-diaphragm; 8) *Anterior-Central Region* (ACR): 6 points (2x3) in the anterior 2 rows and central 3 columns through the hemi-diaphragm; 9) *Posterior-Lateral Region* (PLR): 4 points (2x2) closest to the posterior and lateral aspects of the hemi-diaphragm; 10) *Posterior-Medial Region* (PMR): 4 points (2x2) closest to the posterior and medial aspects of the hemi-diaphragm; 11) *Posterior-Central Region* (PCR): 6 points (2x3) in the posterior 2 rows and central 3 columns through the hemi-diaphragm (see Figure 2(a)); 12) *Central-Lateral Region* (CLR): 6 points (3x2) in the central 3 rows and the 2 columns through the lateral hemi-diaphragm; 13) *Central-Medial Region* (CMR): 6 points (3x2) in the central 3 rows and the 2 columns through the medial hemi-diaphragm.

e-Table 2. Sagittal curvature ($m^{-1}$) of RHD and LHD at end-expiration measured in 13 regions in our subject cohort for each gender. Mean (1st value) and standard deviation (2nd value) are listed.

| **Region** | **Female** | | | **Male** | | | **P value** Female vs. Male | |
|---|---|---|---|---|---|---|---|---|
| | RHD | LHD | P value | RHD | LHD | P value | RHD | LHD |
| AR | 8.25, 1.58 | 5.84, 1.68 | **<0.001** | 8.20, 2.05 | 6.03, 1.99 | **<0.001** | 0.861 | 0.493 |
| PR | 6.42, 1.42 | 9.08, 2.08 | **<0.001** | 6.29, 1.91 | 8.37, 1.97 | **<0.001** | 0.626 | **0.022** |
| LR | 9.90, 1.91 | 10.59, 1.98 | **0.003** | 9.65, 2.01 | 10.49, 1.82 | **0.002** | 0.412 | 0.728 |
| MR | 9.82, 2.33 | 8.83, 2.23 | **<0.001** | 9.51, 2.17 | 8.27, 2.19 | **<0.001** | 0.368 | 0.097 |
| CR | 15.93, 1.89 | 15.49, 2.09 | **0.014** | 15.54, 1.91 | 14.97, 1.81 | **0.005** | 0.178 | 0.079 |
| ALR | 9.16, 2.28 | 7.40, 2.77 | **<0.001** | 9.45, 2.96 | 7.82, 2.49 | **<0.001** | 0.466 | 0.287 |
| AMR | 6.32, 2.64 | 5.04, 2.67 | **<0.001** | 5.99, 2.68 | 4.66, 3.23 | **<0.001** | 0.411 | 0.389 |
| ACR | 9.25, 1.86 | 5.02, 1.74 | **<0.001** | 9.08, 2.09 | 5.33, 2.16 | **<0.001** | 0.580 | 0.308 |
| PLR | 5.82, 2.23 | 8.91, 3.23 | **<0.001** | 5.68, 2.29 | 8.01, 2.17 | **<0.001** | 0.682 | **0.034** |
| PMR | 7.58, 2.71 | 8.33, 3.01 | **0.052** | 7.58, 3.47 | 7.70, 2.93 | 0.796 | 0.992 | 0.158 |
| PCR | 5.70, 1.87 | 9.57, 2.55 | **<0.001** | 5.62, 2.26 | 9.01, 2.54 | **<0.001** | 0.798 | 0.141 |
| CLR | 15.99, 3.08 | 16.59, 2.86 | 0.098 | 15.56, 3.33 | 16.72, 2.88 | **0.006** | 0.377 | 0.755 |
| CMR | 16.11, 3.79 | 14.41, 3.60 | **<0.001** | 15.61, 3.51 | 13.53, 3.56 | **<0.001** | 0.368 | 0.107 |

RHD = Right Hemi-Diaphragm; LHD=Left Hemi-Diaphragm.

1) *Anterior Region* (AR): 10 points (2 (rows) x 5 (columns)) from the 2 (coronally-oriented) rows through the anterior hemi-diaphragm (see Figure 2(a)); 2) *Posterior Region* (PR): 10 points (2x5) from the 2 rows through the posterior hemi-diaphragm; 3) *Lateral Region* (LR): 10 points (5x2) from the 2 (sagittally-oriented) columns through the lateral hemi-diaphragm; 4) *Medial Region* (MR): 10 points (5x2) from the 2 columns through the medial hemi-diaphragm; 5) *Central Region* (CR): 9 points (3x3) in the center of the hemi-diaphragm; 6) *Anterior-Lateral Region* (ALR): 4 points (2x2) closest to the anterior and lateral aspects of the hemi-diaphragm (see Figure 2(a)); 7) *Anterior-Medial Region* (AMR): 4 points (2x2) closest to the anterior and medial aspects of the hemi-diaphragm; 8) *Anterior-Central Region* (ACR): 6 points (2x3) in the anterior 2 rows and central 3 columns through the hemi-diaphragm; 9) *Posterior-Lateral Region* (PLR): 4 points (2x2) closest to the posterior and lateral aspects of the hemi-diaphragm; 10) *Posterior-Medial Region* (PMR): 4 points (2x2) closest to the posterior and medial aspects of the hemi-diaphragm; 11) *Posterior-Central Region* (PCR): 6 points (2x3) in the posterior 2 rows and central 3 columns through the hemi-diaphragm (see Figure 2(a)); 12) *Central-Lateral Region* (CLR): 6 points (3x2) in the central 3 rows and the 2 columns through the lateral hemi-diaphragm; 13) *Central-Medial Region* (CMR): 6 points (3x2) in the central 3 rows and the 2 columns through the medial hemi-diaphragm.

| e-Table 3. Sagittal curvature ($m^{-1}$) of RHD and LHD at end-inspiration measured in 13 regions in our subject cohort for each gender. Mean (1st value) and standard deviation (2nd value) are listed. | | | | | | | | |
|---|---|---|---|---|---|---|---|---|
| **Region** | **Female** | | | **Male** | | | **P value** Female vs. Male | |
| | RHD | LHD | P value | RHD | LHD | P value | RHD | LHD |
| AR | 10.74, 1.95 | 7.63, 2.12 | **<0.001** | 10.56, 2.02 | 7.34, 2.29 | **<0.001** | 0.532 | 0.385 |
| PR | 5.72, 1.37 | 7.58, 1.50 | **<0.001** | 5.52, 1.47 | 7.67, 1.87 | **<0.001** | 0.338 | 0.730 |
| LR | 10.82, 1.84 | 10.29, 2.01 | 0.013 | 10.46, 2.09 | 10.56, 2.12 | 0.684 | 0.225 | 0.390 |
| MR | 10.78, 2.14 | 9.57, 2.07 | **<0.001** | 10.29, 1.83 | 8.94, 1.58 | **<0.001** | 0.106 | **0.025** |
| CR | 16.38, 1.92 | 15.73, 1.85 | **<0.001** | 15.86, 1.82 | 15.11, 1.71 | **<0.001** | **0.069** | **0.023** |
| ALR | 12.01, 3.02 | 8.73, 3.02 | **<0.001** | 11.99, 3.19 | 9.30, 3.36 | **<0.001** | 0.967 | 0.240 |
| AMR | 8.44, 2.66 | 7.05, 3.03 | **<0.001** | 7.96, 3.05 | 5.98, 3.09 | **<0.001** | 0.268 | **0.021** |
| ACR | 11.57, 2.22 | 7.00, 2.21 | **<0.001** | 11.63, 2.12 | 6.80, 2.31 | **<0.001** | 0.848 | 0.545 |
| PLR | 5.94, 2.14 | 7.18, 2.23 | **<0.001** | 5.74, 1.93 | 7.55, 2.50 | **<0.001** | 0.517 | 0.296 |
| PMR | 6.00, 2.35 | 6.91, 2.43 | **0.002** | 5.79, 2.69 | 6.83, 2.68 | **0.005** | 0.580 | 0.850 |
| PCR | 4.63, 1.50 | 8.36, 2.00 | **<0.001** | 4.49, 1.68 | 7.99, 2.12 | **<0.001** | 0.575 | 0.240 |
| CLR | 16.65, 2.46 | 16.24, 2.80 | 0.178 | 16.12, 2.93 | 16.47, 2.82 | 0.295 | 0.195 | 0.581 |
| CMR | 17.68, 3.38 | 15.68, 3.23 | **<0.001** | 16.92, 2.93 | 14.67, 2.52 | **<0.001** | 0.113 | **0.023** |

RHD = Right Hemi-Diaphragm; LHD=Left Hemi-Diaphragm.

1) *Anterior Region* (AR): 10 points (2 (rows) x 5 (columns)) from the 2 (coronally-oriented) rows through the anterior hemi-diaphragm (see Figure 2(a)); 2) *Posterior Region* (PR): 10 points (2x5) from the 2 rows through the posterior hemi-diaphragm; 3) *Lateral Region* (LR): 10 points (5x2) from the 2 (sagittally-oriented) columns through the lateral hemi-diaphragm; 4) *Medial Region* (MR): 10 points (5x2) from the 2 columns through the medial hemi-diaphragm; 5) *Central Region* (CR): 9 points (3x3) in the center of the hemi-diaphragm; 6) *Anterior-Lateral Region* (ALR): 4 points (2x2) closest to the anterior and lateral aspects of the hemi-diaphragm (see Figure 2(a)); 7) *Anterior-Medial Region* (AMR): 4 points (2x2) closest to the anterior and medial aspects of the hemi-diaphragm; 8) *Anterior-Central Region* (ACR): 6 points (2x3) in the anterior 2 rows and central 3 columns through the hemi-diaphragm; 9) *Posterior-Lateral Region* (PLR): 4 points (2x2) closest to the posterior and lateral aspects of the hemi-diaphragm; 10) *Posterior-Medial Region* (PMR): 4 points (2x2) closest to the posterior and medial aspects of the hemi-diaphragm; 11) *Posterior-Central Region* (PCR): 6 points (2x3) in the posterior 2 rows and central 3 columns through the hemi-diaphragm (see Figure 2(a)); 12) *Central-Lateral Region* (CLR): 6 points (3x2) in the central 3 rows and the 2 columns through the lateral hemi-diaphragm; 13) *Central-Medial Region* (CMR): 6 points (3x2) in the central 3 rows and the 2 columns through the medial hemi-diaphragm.

| e-Table 4. Coronal curvature ($m^{-1}$) of RHD and LHD at end-expiration measured in 13 regions in our subject cohort for each gender. Mean (1st value) and standard deviation (2nd value) are listed. | | | | | | | | |
|---|---|---|---|---|---|---|---|---|
| **Region** | **Female** | | | **Male** | | | **P value** Female vs. Male | |
| | RHD | LHD | P value | RHD | LHD | P value | RHD | LHD |
| AR | 3.50, 1.93 | 1.10, 1.67 | **<0.001** | 3.73, 1.70 | 1.09, 1.62 | **<0.001** | 0.402 | 0.960 |
| PR | 4.33, 1.67 | 4.33, 1.64 | 0.999 | 4.09, 1.52 | 3.65, 1.93 | **0.04** | 0.314 | **0.012** |
| LR | 5.19, 2.38 | 5.41, 2.37 | 0.421 | 5.23, 2.51 | 4.91, 2.65 | 0.391 | 0.908 | 0.186 |
| MR | 1.20, 3.50 | -0.15, 2.53 | **0.002** | 1.56, 3.16 | -0.70, 2.78 | **<0.001** | 0.470 | 0.167 |
| CR | 12.11, 3.01 | 8.20, 2.75 | **<0.001** | 11.34, 3.06 | 8.18, 3.14 | **<0.001** | 0.093 | 0.956 |
| ALR | 4.41, 2.46 | 4.62, 3.14 | 0.531 | 4.27, 2.39 | 4.32, 3.01 | 0.895 | 0.701 | 0.523 |
| AMR | 0.43, 4.62 | -0.87, 2.95 | **0.019** | 1.53, 4.33 | -0.92, 3.41 | **<0.001** | 0.105 | 0.910 |
| ACR | 5.62, 3.06 | 1.30, 2.66 | **<0.001** | 5.92, 3.04 | 1.57, 2.59 | **<0.001** | 0.505 | 0.508 |
| PLR | 4.43, 2.95 | 4.26, 2.33 | 0.628 | 4.78, 2.75 | 3.76, 2.77 | **0.009** | 0.410 | 0.198 |
| PMR | 0.92, 2.92 | 0.75, 3.09 | 0.684 | 0.72, 2.93 | -0.19, 3.46 | **0.035** | 0.648 | **0.057** |
| PCR | 7.19, 2.58 | 7.43, 2.75 | 0.468 | 6.63, 2.47 | 6.62, 2.93 | 0.976 | 0.146 | **0.059** |
| CLR | 8.25, 2.98 | 8.83, 3.53 | 0.152 | 8.33, 3.86 | 8.34, 3.82 | 0.987 | 0.876 | 0.374 |
| CMR | 2.25, 4.70 | -0.15, 3.74 | **<0.001** | 2.28, 4.37 | -1.07, 4.30 | **<0.001** | 0.970 | 0.130 |

RHD = Right Hemi-Diaphragm; LHD=Left Hemi-Diaphragm.

1) *Anterior Region* (AR): 10 points (2 (rows) x 5 (columns)) from the 2 (coronally-oriented) rows through the anterior hemi-diaphragm (see Figure 2(a)); 2) *Posterior Region* (PR): 10 points (2x5) from the 2 rows through the posterior hemi-diaphragm; 3) *Lateral Region* (LR): 10 points (5x2) from the 2 (sagittally-oriented) columns through the lateral hemi-diaphragm; 4) *Medial Region* (MR): 10 points (5x2) from the 2 columns through the medial hemi-diaphragm; 5) *Central Region* (CR): 9 points (3x3) in the center of the hemi-diaphragm; 6) *Anterior-Lateral Region* (ALR): 4 points (2x2) closest to the anterior and lateral aspects of the hemi-diaphragm (see Figure 2(a)); 7) *Anterior-Medial Region* (AMR): 4 points (2x2) closest to the anterior and medial aspects of the hemi-diaphragm; 8) *Anterior-Central Region* (ACR): 6 points (2x3) in the anterior 2 rows and central 3 columns through the hemi-diaphragm; 9) *Posterior-Lateral Region* (PLR): 4 points (2x2) closest to the posterior and lateral aspects of the hemi-diaphragm; 10) *Posterior-Medial Region* (PMR): 4 points (2x2) closest to the posterior and medial aspects of the hemi-diaphragm; 11) *Posterior-Central Region* (PCR): 6 points (2x3) in the posterior 2 rows and central 3 columns through the hemi-diaphragm (see Figure 2(a)); 12) *Central-Lateral Region* (CLR): 6 points (3x2) in the central 3 rows and the 2 columns through the lateral hemi-diaphragm; 13) *Central-Medial Region* (CMR): 6 points (3x2) in the central 3 rows and the 2 columns through the medial hemi-diaphragm.

e-Table 5. Coronal curvature ($m^{-1}$) of RHD and LHD at end-inspiration measured in 13 regions in our subject cohort for each gender. Mean (1st value) and standard deviation (2nd value) are listed.

| **Region** | **Female** | | | **Male** | | | **P value** Female vs. Male | |
|---|---|---|---|---|---|---|---|---|
| | RHD | LHD | P value | RHD | LHD | P value | RHD | LHD |
| AR | 3.69, 2.07 | 0.89, 2.13 | **<0.001** | 3.51, 2.01 | 0.79, 1.80 | **<0.001** | 0.564 | 0.732 |
| PR | 4.10, 2.34 | 3.61, 2.03 | 0.116 | 3.75, 2.47 | 3.58, 2.26 | 0.625 | 0.336 | 0.929 |
| LR | 6.31, 3.05 | 4.85, 2.20 | **<0.001** | 5.43, 4.10 | 4.64, 2.88 | 0.143 | 0.106 | 0.592 |
| MR | 0.90, 3.37 | -0.61, 2.91 | **0.001** | 0.35, 3.29 | -0.88, 3.15 | **0.012** | 0.273 | 0.564 |
| CR | 11.69, 3.55 | 7.71, 3.37 | **<0.001** | 10.25, 4.28 | 7.13, 3.16 | **<0.001** | **0.015** | 0.243 |
| ALR | 5.44, 2.86 | 4.71, 2.71 | **0.079** | 4.74, 3.59 | 4.63, 2.52 | 0.813 | 0.153 | 0.837 |
| AMR | -0.05, 4.64 | -1.02, 4.05 | 0.102 | -0.32, 4.53 | -1.01, 4.15 | 0.287 | 0.699 | 0.985 |
| ACR | 5.64, 3.35 | 0.85, 3.55 | **<0.001** | 5.30, 3.43 | 0.61, 3.11 | **<0.001** | 0.501 | 0.626 |
| PLR | 5.56, 3.56 | 3.56, 2.51 | **<0.001** | 4.92, 5.28 | 3.44, 3.82 | **0.028** | 0.341 | 0.805 |
| PMR | 0.58, 3.11 | -0.13, 3.48 | 0.153 | -0.22, 3.27 | -0.40, 3.44 | 0.723 | 0.099 | 0.610 |
| PCR | 6.73, 3.87 | 6.22, 3.42 | 0.324 | 6.06, 4.11 | 5.91, 3.81 | 0.785 | 0.262 | 0.567 |
| CLR | 9.80, 3.97 | 7.69, 3.38 | **<0.001** | 8.38, 4.94 | 7.34, 4.00 | 0.135 | **0.036** | 0.537 |
| CMR | 2.01, 4.61 | -0.84, 3.96 | **<0.001** | 1.25, 4.40 | -1.13, 4.20 | **0.001** | 0.267 | 0.634 |

RHD = Right Hemi-Diaphragm; LHD=Left Hemi-Diaphragm.

1) *Anterior Region* (AR): 10 points (2 (rows) x 5 (columns)) from the 2 (coronally-oriented) rows through the anterior hemi-diaphragm (see Figure 2(a)); 2) *Posterior Region* (PR): 10 points (2x5) from the 2 rows through the posterior hemi-diaphragm; 3) *Lateral Region* (LR): 10 points (5x2) from the 2 (sagittally-oriented) columns through the lateral hemi-diaphragm; 4) *Medial Region* (MR): 10 points (5x2) from the 2 columns through the medial hemi-diaphragm; 5) *Central Region* (CR): 9 points (3x3) in the center of the hemi-diaphragm; 6) *Anterior-Lateral Region* (ALR): 4 points (2x2) closest to the anterior and lateral aspects of the hemi-diaphragm (see Figure 2(a)); 7) *Anterior-Medial Region* (AMR): 4 points (2x2) closest to the anterior and medial aspects of the hemi-diaphragm; 8) *Anterior-Central Region* (ACR): 6 points (2x3) in the anterior 2 rows and central 3 columns through the hemi-diaphragm; 9) *Posterior-Lateral Region* (PLR): 4 points (2x2) closest to the posterior and lateral aspects of the hemi-diaphragm; 10) *Posterior-Medial Region* (PMR): 4 points (2x2) closest to the posterior and medial aspects of the hemi-diaphragm; 11) *Posterior-Central Region* (PCR): 6 points (2x3) in the posterior 2 rows and central 3 columns through the hemi-diaphragm (see Figure 2(a)); 12) *Central-Lateral Region* (CLR): 6 points (3x2) in the central 3 rows and the 2 columns through the lateral hemi-diaphragm; 13) *Central-Medial Region* (CMR): 6 points (3x2) in the central 3 rows and the 2 columns through the medial hemi-diaphragm.

e-Table 6. Pearson correlations in velocity between right hemi-diaphragm (RHD) and left hemi-diaphragm (LHD) in 13 regions in our subject cohort for each gender and age group. Age ranges are indicated by half open intervals. For example, [6, 8) = 6 ≤ age < 8. Regions showing little or very weak correlations (<0.2 in magnitude) are highlighted.

| **Female** | | | | | |
|---|---|---|---|---|---|
| Region | [6, 8) | [8, 10) | [10, 12) | [12, 14) | [14, 20) |
| AR | 0.75 | 0.88 | 0.84 | 0.75 | 0.49 |
| PR | 0.94 | 0.75 | 0.25 | 0.78 | 0.18 |
| LR | 0.77 | 0.44 | 0.41 | 0.78 | 0.57 |
| MR | 0.33 | 0.92 | 0.29 | 0.56 | 0.35 |
| CR | 0.85 | 0.8 | -0.05 | 0.71 | 0.36 |
| ALR | 0.72 | 0.44 | 0.48 | 0.79 | 0.55 |
| AMR | 0.27 | 0.92 | 0.77 | -0.14 | 0.4 |
| ACR | 0.36 | 0.84 | 0.48 | 0.77 | 0.04 |
| PLR | 0.83 | 0.42 | 0.16 | 0.56 | 0.48 |
| PMR | 0.46 | 0.89 | -0.06 | 0.7 | 0.1 |
| PCR | 0.89 | 0.67 | -0.46 | 0.62 | 0.12 |
| CLR | 0.74 | 0.44 | 0.27 | 0.88 | 0.6 |
| CMR | 0.33 | 0.9 | 0.11 | 0.49 | 0.24 |
| **Male** | | | | | |
| Region | [6, 8) | [8, 10) | [10, 12) | [12, 14) | [14, 20) |
| AR | 0.21 | 0.36 | 0.78 | 0.82 | -0.04 |
| PR | 0.56 | 0.59 | 0.5 | 0.46 | 0.39 |
| LR | 0.18 | 0.4 | 0.32 | 0.38 | -0.08 |
| MR | 0.29 | 0.54 | 0.55 | 0.41 | 0.41 |
| CR | 0.54 | 0.6 | 0.67 | 0.17 | -0.23 |
| ALR | 0.4 | 0.28 | 0.04 | 0.59 | -0.31 |
| AMR | 0.01 | 0.61 | 0.6 | 0.85 | 0.42 |
| ACR | 0.5 | 0.48 | 0.86 | 0.73 | -0.44 |
| PLR | 0.18 | 0.33 | 0.39 | 0.27 | 0.21 |
| PMR | 0.66 | 0.28 | 0.52 | 0.16 | 0.36 |
| PCR | 0.76 | 0.7 | 0.54 | 0.52 | 0.23 |
| CLR | 0.19 | 0.29 | 0.28 | 0.42 | -0.11 |
| CMR | -0.02 | 0.62 | 0.58 | 0.27 | 0.31 |

RHD = Right Hemi-Diaphragm; LHD=Left Hemi-Diaphragm.

1) *Anterior Region* (AR): 10 points (2 (rows) x 5 (columns)) from the 2 (coronally-oriented) rows through the anterior hemi-diaphragm (see Figure 2(a)); 2) *Posterior Region* (PR): 10 points (2x5) from the 2 rows through the posterior hemi-diaphragm; 3) *Lateral Region* (LR): 10 points (5x2) from the 2 (sagittally-oriented) columns through the lateral hemi-diaphragm; 4) *Medial Region* (MR): 10 points (5x2) from the 2 columns through the medial hemi-diaphragm; 5) *Central Region* (CR): 9 points (3x3) in the center of the hemi-diaphragm; 6) *Anterior-Lateral Region* (ALR): 4 points (2x2) closest to the anterior and lateral aspects of the hemi-diaphragm (see Figure 2(a)); 7) *Anterior-Medial Region* (AMR): 4 points (2x2) closest to the anterior and medial aspects of the hemi-diaphragm; 8) *Anterior-Central Region* (ACR): 6 points (2x3) in the anterior 2 rows and central 3 columns through the hemi-diaphragm; 9) *Posterior-Lateral Region* (PLR): 4 points (2x2) closest to the posterior and lateral aspects of the hemi-diaphragm; 10) *Posterior-Medial Region* (PMR): 4 points (2x2) closest to the posterior and medial aspects of the hemi-diaphragm; 11) *Posterior-Central Region* (PCR): 6 points (2x3) in the posterior 2 rows and central 3 columns through the hemi-diaphragm (see Figure 2(a)); 12) *Central-Lateral Region* (CLR): 6 points (3x2) in the central 3 rows and the 2 columns through the lateral hemi-diaphragm; 13) *Central-Medial Region* (CMR): 6 points (3x2) in the central 3 rows and the 2 columns through the medial hemi-diaphragm.

e-Table 7. Pearson correlation between velocity and change in sagittal curvature from end-expiration to end-inspiration of RHD and LHD measured in 13 regions in our subject cohort for each gender and age group. Age ranges are indicated by half open intervals. For example, [6, 8) = 6 $\leq$ age < 8. Moderate to high magnitudes of correlations ($\geq$0.5) are highlighted.

| **Female** | | | | | | | | | | |
|---|---|---|---|---|---|---|---|---|---|---|
| Region | RHD | | | | | LHD | | | | |
| | [6, 8) | [8, 10) | [10, 12) | [12, 14) | [14, 20) | [6, 8) | [8, 10) | [10, 12) | [12, 14) | [14, 20) |
| AR | 0.06 | 0.37 | -0.2 | -0.2 | -0.3 | -0.5 | -0.33 | **-0.55** | -0.22 | -0.04 |
| PR | 0.39 | 0.31 | 0.23 | 0.48 | -0.14 | 0.19 | 0.09 | -0.22 | 0.26 | 0.33 |
| LR | 0.04 | 0.19 | 0 | 0.01 | -0.19 | -0.36 | -0.07 | -0.31 | 0.21 | -0.43 |
| MR | -0.45 | -0.01 | 0.02 | 0.26 | 0.02 | -0.08 | -0.2 | -0.43 | -0.13 | 0.13 |
| CR | 0.45 | 0.15 | 0.34 | **0.55** | 0.08 | 0.32 | 0.34 | -0.08 | 0.45 | 0.2 |
| ALR | -0.01 | -0.08 | 0.4 | 0.06 | -0.28 | -0.43 | -0.21 | -0.45 | 0.2 | -0.23 |
| AMR | -0.46 | -0.04 | -0.2 | -0.03 | -0.22 | -0.14 | -0.4 | -0.43 | 0.22 | 0.23 |
| ACR | -0.18 | 0.24 | -0.21 | 0.04 | -0.09 | 0.06 | 0.01 | -0.45 | 0.07 | 0.07 |
| PLR | 0.14 | 0.37 | -0.23 | 0.15 | 0.08 | -0.2 | 0.14 | -0.43 | 0.22 | 0.19 |
| PMR | 0.08 | 0.05 | -0.01 | **0.54** | 0.09 | 0.6 | -0.14 | -0.02 | 0.49 | 0.3 |
| PCR | **0.56** | 0.31 | **0.56** | 0.39 | -0.2 | 0.12 | 0.23 | 0.45 | **0.79** | 0.44 |
| CLR | 0.05 | 0.32 | 0.24 | 0.31 | 0.18 | -0.25 | -0.04 | -0.08 | 0.36 | -0.18 |
| CMR | -0.25 | 0.21 | 0.34 | 0.44 | 0.45 | 0.04 | -0.02 | -0.07 | 0.34 | 0.32 |
| **Male** | | | | | | | | | | |
| Region | RHD | | | | | LHD | | | | |
| | [6, 8) | [8, 10) | [10, 12) | [12, 14) | [14, 20) | [6, 8) | [8, 10) | [10, 12) | [12, 14) | [14, 20) |
| AR | 0.03 | -0.14 | -0.21 | **-0.55** | 0.15 | -0.01 | **-0.52** | -0.4 | **-0.57** | -0.3 |
| PR | 0.13 | 0.38 | 0.51 | **0.86** | 0.03 | -0.42 | 0.28 | 0.22 | 0.16 | 0.26 |
| LR | -0.31 | 0.13 | 0.21 | -0.01 | -0.31 | -0.22 | -0.12 | -0.35 | -0.31 | -0.02 |
| MR | -0.4 | 0.01 | 0.14 | 0.15 | 0.17 | 0.29 | -0.36 | -0.34 | 0.32 | 0.07 |
| CR | 0.24 | 0.12 | 0.13 | -0.11 | -0.08 | -0.17 | **0.73** | -0.01 | 0.18 | 0.25 |
| ALR | -0.14 | 0.15 | -0.18 | -0.09 | 0.05 | 0.16 | -0.26 | **-0.58** | -0.29 | -0.1 |
| AMR | -0.01 | 0.05 | 0.05 | -0.37 | 0.07 | 0.42 | -0.15 | -0.27 | -0.46 | -0.14 |
| ACR | -0.09 | -0.33 | **0.53** | **-0.53** | -0.04 | -0.13 | -0.07 | -0.13 | -0.17 | -0.29 |
| PLR | **0.61** | 0.35 | 0.4 | **0.73** | -0.18 | **-0.65** | 0.12 | -0.24 | -0.37 | 0.24 |
| PMR | -0.09 | 0.23 | **0.59** | **0.67** | 0.19 | 0.47 | -0.11 | -0.16 | **0.61** | 0.16 |
| PCR | 0.08 | **0.64** | **0.51** | 0.1 | 0.16 | 0.34 | 0.37 | 0.28 | -0.28 | 0.26 |
| CLR | -0.24 | 0.31 | 0.34 | 0.1 | -0.26 | -0.11 | 0.24 | -0.07 | -0.09 | 0.11 |
| CMR | -0.07 | 0.4 | 0.34 | 0.39 | 0.35 | **0.51** | -0.14 | 0.15 | **0.61** | 0.46 |

RHD = Right Hemi-Diaphragm; LHD=Left Hemi-Diaphragm.

1) *Anterior Region* (AR): 10 points (2 (rows) x 5 (columns)) from the 2 (coronally-oriented) rows through the anterior hemi-diaphragm (see Figure 2(a)); 2) *Posterior Region* (PR): 10 points (2x5) from the 2 rows through the posterior hemi-diaphragm; 3) *Lateral Region* (LR): 10 points (5x2) from the 2 (sagittally-oriented) columns through the lateral hemi-diaphragm; 4) *Medial Region* (MR): 10 points (5x2) from the 2 columns through the medial hemi-diaphragm; 5) *Central Region* (CR): 9 points (3x3) in the center of the hemi-diaphragm; 6) *Anterior-Lateral Region* (ALR): 4 points (2x2) closest to the anterior and lateral aspects of the hemi-diaphragm (see Figure 2(a)); 7) *Anterior-Medial Region* (AMR): 4 points (2x2) closest to the anterior and medial aspects of the hemi-diaphragm; 8) *Anterior-Central Region* (ACR): 6 points (2x3) in the anterior 2 rows and central 3 columns through the hemi-diaphragm; 9) *Posterior-Lateral Region* (PLR): 4 points (2x2) closest to the posterior and lateral aspects of the hemi-diaphragm; 10) *Posterior-Medial Region* (PMR): 4 points (2x2) closest to the

posterior and medial aspects of the hemi-diaphragm; 11) *Posterior-Central Region* (PCR): 6 points (2x3) in the posterior 2 rows and central 3 columns through the hemi-diaphragm (see Figure 2(a)); 12) *Central-Lateral Region* (CLR): 6 points (3x2) in the central 3 rows and the 2 columns through the lateral hemi-diaphragm; 13) *Central-Medial Region* (CMR): 6 points (3x2) in the central 3 rows and the 2 columns through the medial hemi-diaphragm.

e-Table 8. Pearson correlation between velocity and change in coronal curvature from end-expiration to end-inspiration of RHD and LHD measured in 13 regions in our subject cohort for each gender and age group. Age ranges are indicated by half open intervals. For example, [6, 8) = 6 ≤ age < 8. Moderate to high magnitudes of correlations (≥0.5) are highlighted.

| **Female** | | | | | | | | | | |
|---|---|---|---|---|---|---|---|---|---|---|
| Region | RHD | | | | | LHD | | | | |
| | [6, 8) | [8, 10) | [10, 12) | [12, 14) | [14, 20) | [6, 8) | [8, 10) | [10, 12) | [12, 14) | [14, 20) |
| AR | -0.24 | -0.1 | -0.22 | 0.12 | -0.08 | -0.19 | -0.18 | 0.07 | -0.12 | -0.07 |
| PR | -0.11 | -0.23 | 0.1 | 0 | 0.43 | 0.62 | -0.07 | 0.22 | 0.22 | 0.62 |
| LR | **-0.61** | -0.01 | 0.2 | 0.1 | 0.33 | -0.26 | -0.05 | -0.4 | 0.05 | 0.09 |
| MR | 0.35 | 0.28 | 0.34 | -0.11 | 0.07 | 0.15 | 0.22 | 0.36 | 0.16 | 0.22 |
| CR | 0.04 | -0.18 | **0.67** | -0.17 | 0.37 | 0.51 | 0.07 | 0.39 | 0.19 | 0.47 |
| ALR | **-0.55** | 0.05 | 0.49 | 0.19 | 0.28 | -0.17 | -0.24 | **-0.56** | 0.42 | 0.06 |
| AMR | 0.2 | 0.29 | -0.08 | 0.15 | 0.04 | -0.03 | -0.1 | 0.45 | 0.07 | -0.14 |
| ACR | 0.37 | 0.03 | 0.2 | 0.23 | 0.11 | -0.05 | -0.06 | 0.37 | -0.12 | 0.2 |
| PLR | -0.49 | 0.07 | 0.29 | 0.04 | 0.42 | -0.42 | -0.04 | -0.13 | -0.13 | 0.32 |
| PMR | 0.29 | 0.02 | 0.05 | 0.11 | 0.07 | 0.49 | -0.11 | 0.16 | 0.44 | 0.39 |
| PCR | 0.27 | -0.05 | **0.62** | 0 | 0.43 | **0.65** | 0.07 | 0.16 | **0.66** | **0.64** |
| CLR | **-0.66** | -0.24 | 0.03 | -0.06 | 0.23 | -0.17 | 0.06 | -0.51 | -0.08 | 0.04 |
| CMR | 0.23 | 0.31 | 0.19 | -0.21 | 0.21 | 0.42 | 0.16 | **0.63** | -0.02 | 0.42 |
| **Male** | | | | | | | | | | |
| Region | RHD | | | | | LHD | | | | |
| | [6, 8) | [8, 10) | [10, 12) | [12, 14) | [14, 20) | [6, 8) | [8, 10) | [10, 12) | [12, 14) | [14, 20) |
| AR | 0.35 | 0.19 | 0 | 0.07 | -0.01 | 0 | -0.16 | -0.38 | -0.19 | 0.12 |
| PR | -0.26 | 0.02 | 0.7 | **0.61** | 0.34 | 0.5 | 0.02 | -0.01 | **0.74** | 0.15 |
| LR | 0.12 | -0.25 | 0.49 | 0.37 | 0.2 | **-0.58** | -0.27 | 0.44 | 0.48 | 0.26 |
| MR | 0.17 | 0 | 0.36 | 0.05 | 0.08 | 0.32 | 0.47 | -0.36 | 0.02 | 0.25 |
| CR | **0.54** | 0.09 | **0.76** | 0.6 | 0.36 | 0.27 | **0.69** | **0.51** | 0.49 | 0.45 |
| ALR | 0.3 | 0.01 | 0.19 | **0.51** | 0.27 | -0.28 | -0.38 | 0 | 0.19 | 0.38 |
| AMR | 0.32 | -0.26 | 0.37 | -0.22 | -0.09 | 0.44 | 0.42 | -0.36 | 0.16 | -0.06 |
| ACR | **0.52** | 0.11 | **0.56** | 0.13 | 0.3 | 0.33 | 0.07 | -0.12 | -0.01 | 0.3 |
| PLR | 0.22 | -0.41 | **0.67** | 0.38 | 0.05 | -0.36 | 0.05 | 0.26 | **0.58** | 0.2 |
| PMR | -0.11 | 0.03 | 0.46 | 0.35 | 0.43 | 0.34 | 0.33 | 0.07 | 0.35 | 0.16 |
| PCR | -0.1 | 0.26 | **0.78** | **0.69** | 0.46 | **0.67** | **0.5** | 0.08 | **0.64** | 0.38 |
| CLR | 0.19 | -0.06 | 0.44 | 0.01 | 0.21 | **-0.52** | -0.44 | 0.6 | 0.32 | 0.21 |
| CMR | 0.09 | 0.05 | **0.53** | 0.27 | 0.23 | 0.21 | 0.37 | -0.03 | -0.06 | **0.54** |

RHD = Right Hemi-Diaphragm; LHD=Left Hemi-Diaphragm.

1) *Anterior Region* (AR): 10 points (2 (rows) x 5 (columns)) from the 2 (coronally-oriented) rows through the anterior hemi-diaphragm (see Figure 2(a)); 2) *Posterior Region* (PR): 10 points (2x5) from the 2 rows through the posterior hemi-diaphragm; 3) *Lateral Region* (LR): 10 points (5x2) from the 2 (sagittally-oriented) columns

through the lateral hemi-diaphragm; 4) *Medial Region* (MR): 10 points (5x2) from the 2 columns through the medial hemi-diaphragm; 5) *Central Region* (CR): 9 points (3x3) in the center of the hemi-diaphragm; 6) *Anterior-Lateral Region* (ALR): 4 points (2x2) closest to the anterior and lateral aspects of the hemi-diaphragm (see Figure 2(a)); 7) *Anterior-Medial Region* (AMR): 4 points (2x2) closest to the anterior and medial aspects of the hemi-diaphragm; 8) *Anterior-Central Region* (ACR): 6 points (2x3) in the anterior 2 rows and central 3 columns through the hemi-diaphragm; 9) *Posterior-Lateral Region* (PLR): 4 points (2x2) closest to the posterior and lateral aspects of the hemi-diaphragm; 10) *Posterior-Medial Region* (PMR): 4 points (2x2) closest to the posterior and medial aspects of the hemi-diaphragm; 11) *Posterior-Central Region* (PCR): 6 points (2x3) in the posterior 2 rows and central 3 columns through the hemi-diaphragm (see Figure 2(a)); 12) *Central-Lateral Region* (CLR): 6 points (3x2) in the central 3 rows and the 2 columns through the lateral hemi-diaphragm; 13) *Central-Medial Region* (CMR): 6 points (3x2) in the central 3 rows and the 2 columns through the medial hemi-diaphragm.

e-Table 9. Velocity (mm/s) of RHD and LHD measured in 13 regions for a male TIS patient pre-operatively and post-operatively and for their matched normal male subjects, respectively.

| Region | **Pre-op** | | **Post-op** | | **Normal** | | **Normal** | |
|---|---|---|---|---|---|---|---|---|
| | RHD | LHD | RHD | LHD | RHD | LHD | RHD | LHD |
| AR | 4.13 | 2.87 | 5.67 | 5.00 | 8.39 | 5.49 | 3.76 | 1.42 |
| PR | 4.83 | 3.57 | 8.58 | 8.33 | 8.85 | 8.09 | 7.53 | 7.22 |
| LR | 6.51 | 2.80 | 8.50 | 6.00 | 7.48 | 4.88 | 3.66 | 4.27 |
| MR | 2.73 | 3.64 | 5.58 | 8.33 | 8.54 | 7.93 | 5.49 | 3.26 |
| CR | 3.58 | 3.81 | 7.59 | 6.67 | 10.85 | 7.97 | 6.10 | 4.63 |
| ALR | 6.13 | 2.10 | 6.88 | 4.58 | 8.01 | 2.67 | 3.05 | 1.27 |
| AMR | 2.80 | 3.50 | 5.42 | 8.75 | 8.01 | 8.39 | 3.05 | 1.27 |
| ACR | 3.50 | 3.03 | 4.86 | 3.33 | 8.65 | 7.88 | 4.07 | 1.53 |
| PLR | 7.18 | 3.15 | 10.21 | 8.75 | 6.87 | 7.63 | 4.32 | 8.90 |
| PMR | 2.80 | 4.03 | 6.04 | 7.71 | 9.16 | 7.25 | 8.90 | 5.09 |
| PCR | 4.90 | 3.97 | 9.31 | 8.19 | 10.68 | 9.92 | 8.99 | 7.80 |
| CLR | 6.07 | 3.15 | 8.75 | 5.42 | 7.63 | 4.32 | 3.39 | 2.88 |
| CMR | 2.68 | 3.50 | 5.28 | 8.33 | 8.39 | 8.14 | 4.41 | 3.90 |

RHD = Right Hemi-Diaphragm; LHD=Left Hemi-Diaphragm.

1) *Anterior Region* (AR): 10 points (2 (rows) x 5 (columns)) from the 2 (coronally-oriented) rows through the anterior hemi-diaphragm (see Figure 2(a)); 2) *Posterior Region* (PR): 10 points (2x5) from the 2 rows through the posterior hemi-diaphragm; 3) *Lateral Region* (LR): 10 points (5x2) from the 2 (sagittally-oriented) columns through the lateral hemi-diaphragm; 4) *Medial Region* (MR): 10 points (5x2) from the 2 columns through the medial hemi-diaphragm; 5) *Central Region* (CR): 9 points (3x3) in the center of the hemi-diaphragm; 6) *Anterior-Lateral Region* (ALR): 4 points (2x2) closest to the anterior and lateral aspects of the hemi-diaphragm (see Figure 2(a)); 7) *Anterior-Medial Region* (AMR): 4 points (2x2) closest to the anterior and medial aspects of the hemi-diaphragm; 8) *Anterior-Central Region* (ACR): 6 points (2x3) in the anterior 2 rows and central 3 columns through the hemi-diaphragm; 9) *Posterior-Lateral Region* (PLR): 4 points (2x2) closest to the posterior and lateral aspects of the hemi-diaphragm; 10) *Posterior-Medial Region* (PMR): 4 points (2x2) closest to the posterior and medial aspects of the hemi-diaphragm; 11) *Posterior-Central Region* (PCR): 6 points (2x3) in the posterior 2 rows and central 3 columns through the hemi-diaphragm (see Figure 2(a)); 12) *Central-Lateral Region* (CLR): 6 points (3x2) in the central 3 rows and the 2 columns through the lateral hemi-diaphragm; 13) *Central-Medial Region* (CMR): 6 points (3x2) in the central 3 rows and the 2 columns through the medial hemi-diaphragm.

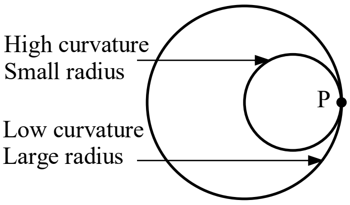


e-Figure 1. Examples of curves with high and low curvatures at point P.

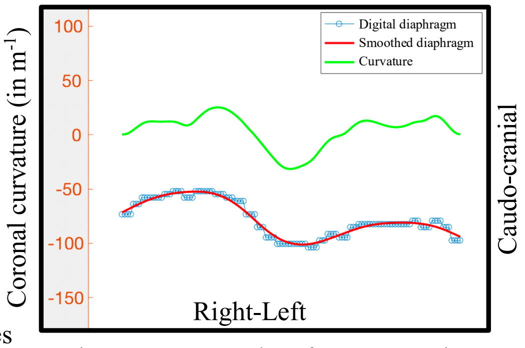


e-Figure 2. Example of curvature in a sagittal plane passing through the right hemi-diaphragm of a normal subject at end-expiration. The digital sagittal curve, the smoothed version of the digital sagittal curve, and the sagittal curvature shown as a graph are displayed.

e-Figure 3. Plots of mean velocity in each region for the different age groups and right hemi-diaphragm (RHD) and left hemi-diaphragm (LHD) for both genders. Mean ± one standard deviation limits are also shown.

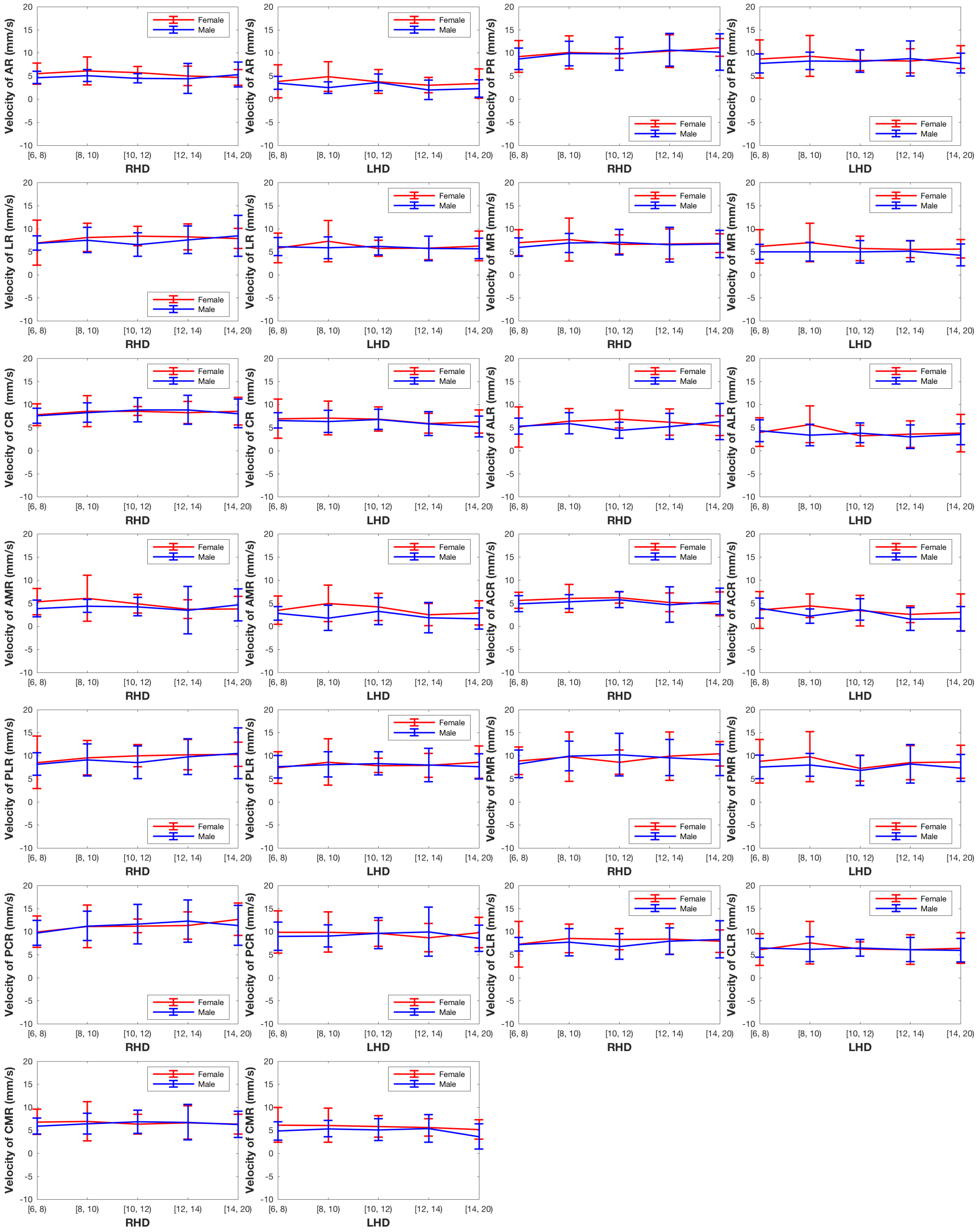

e-Figure 4. Plots of mean sagittal curvature in each region for the different age groups and right hemi-diaphragm (RHD) and left hemi-diaphragm (LHD) at end-expiration (EE) for both genders. Mean $\pm$ one standard deviation limits are also shown.

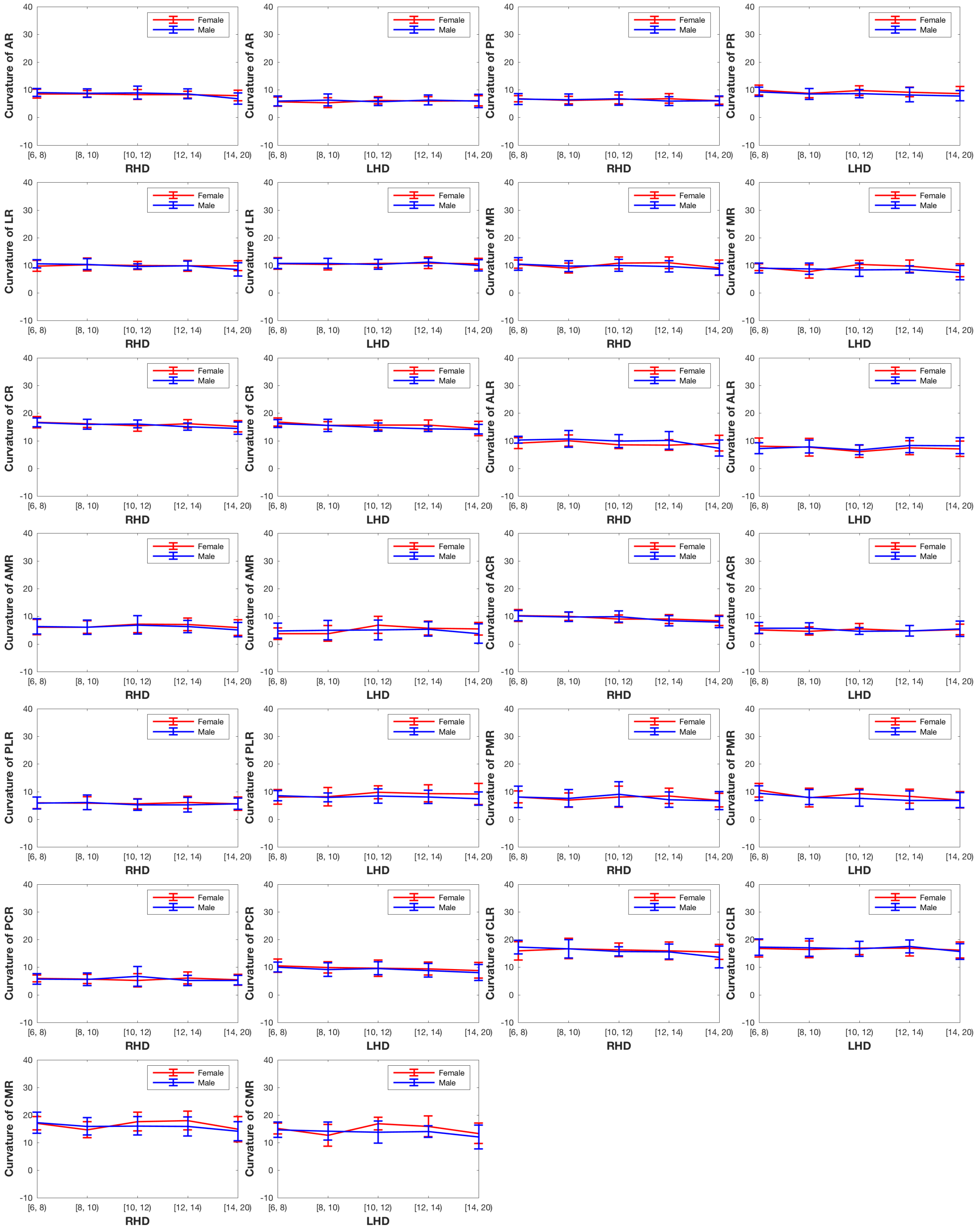

e-Figure 5. Plots of mean sagittal curvature in each region for the different age groups and right hemi-diaphragm (RHD) and left hemi-diaphragm (LHD) at end-inspiration (EI) for both genders. Mean ± one standard deviation limits are also shown.

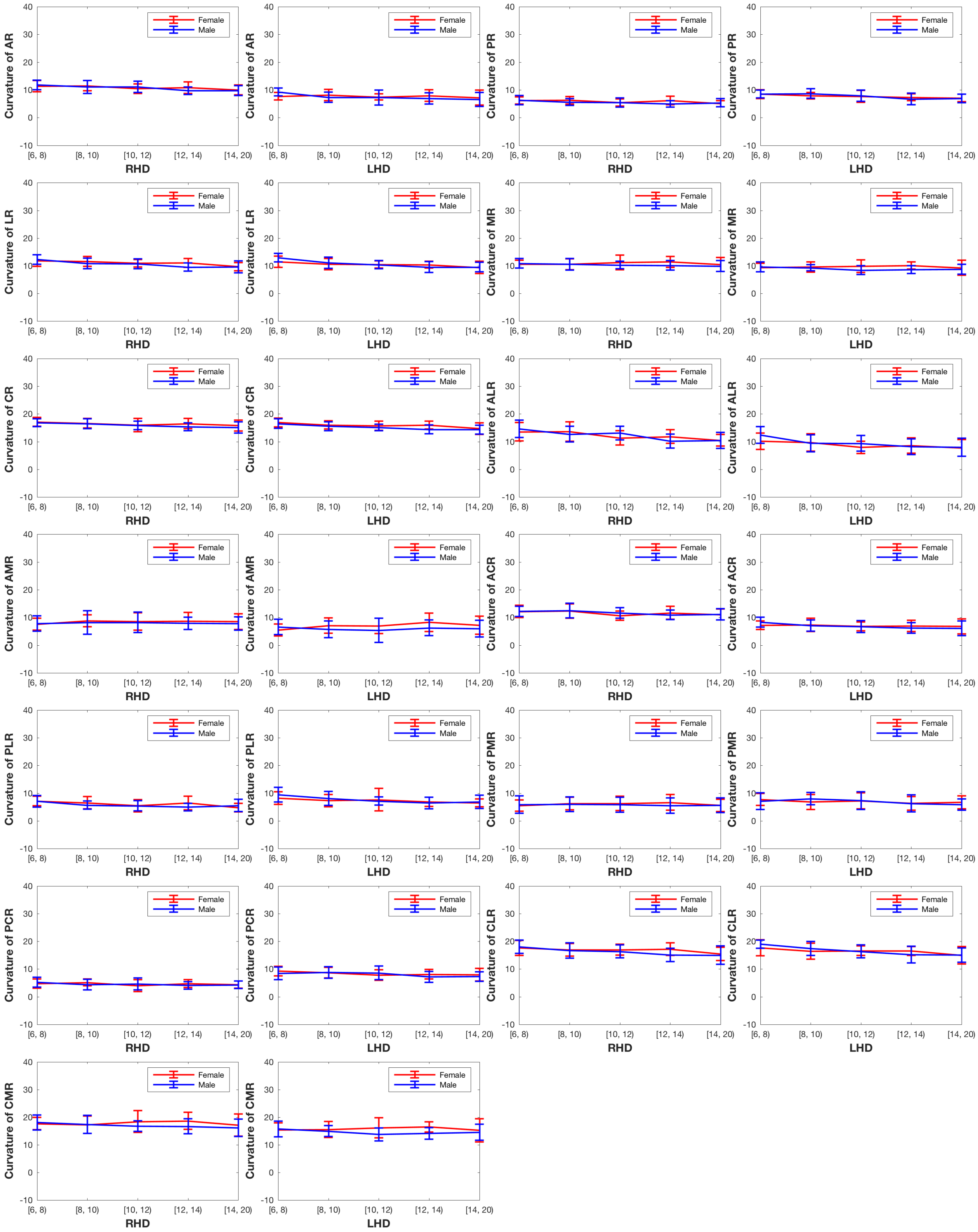

e-Figure 6. Plots of mean coronal curvature in each region for the different age groups and right hemi-diaphragm (RHD) and left hemi-diaphragm (LHD) at end-expiration (EE) for both genders. Mean $\pm$ one standard deviation limits are also shown.

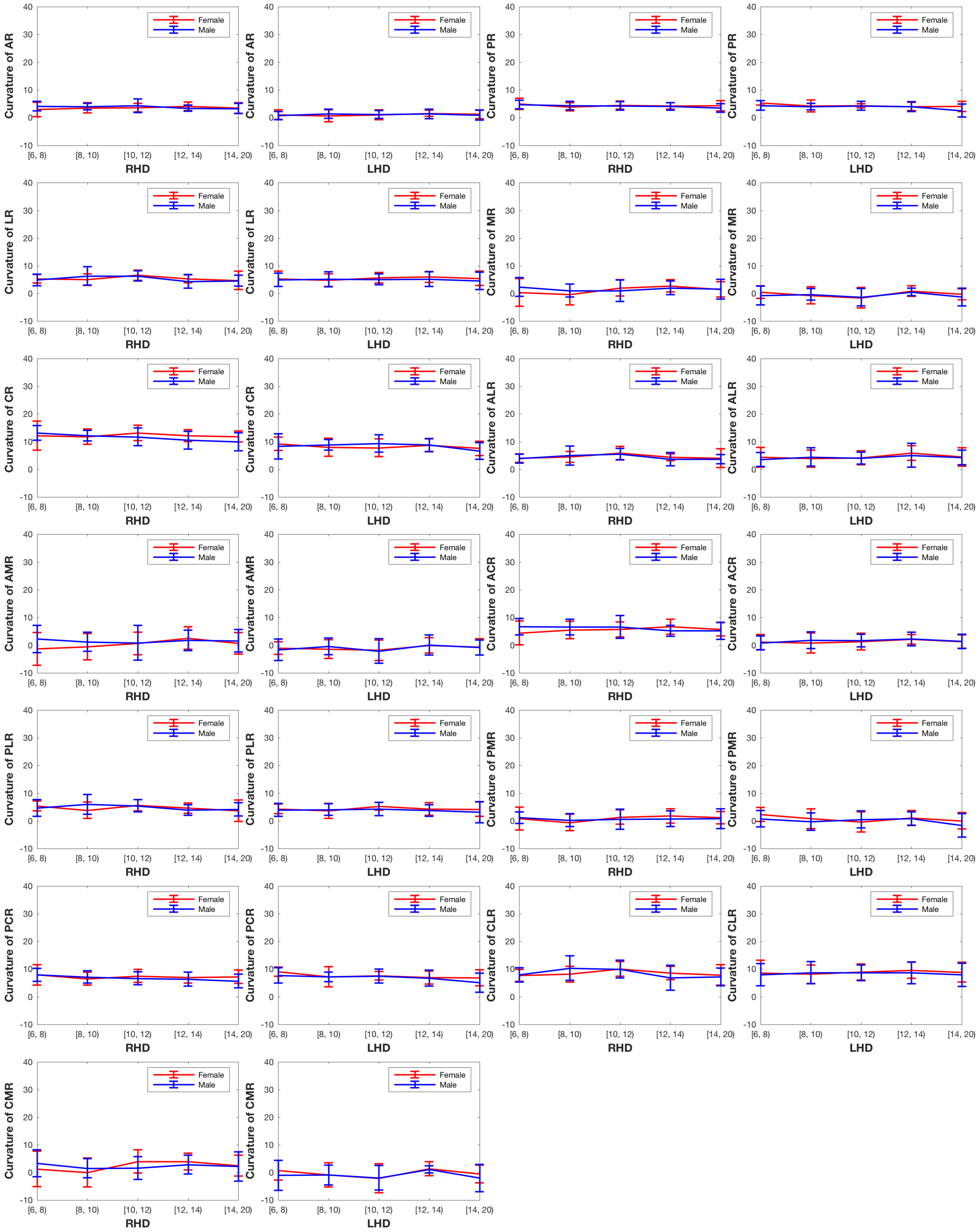

e-Figure 7. Plots of mean coronal curvature in each region for the different age groups and right hemi-diaphragm (RHD) and left hemi-diaphragm (LHD) at end-inspiration (EI) for both genders. Mean ± one standard deviation limits are also shown.

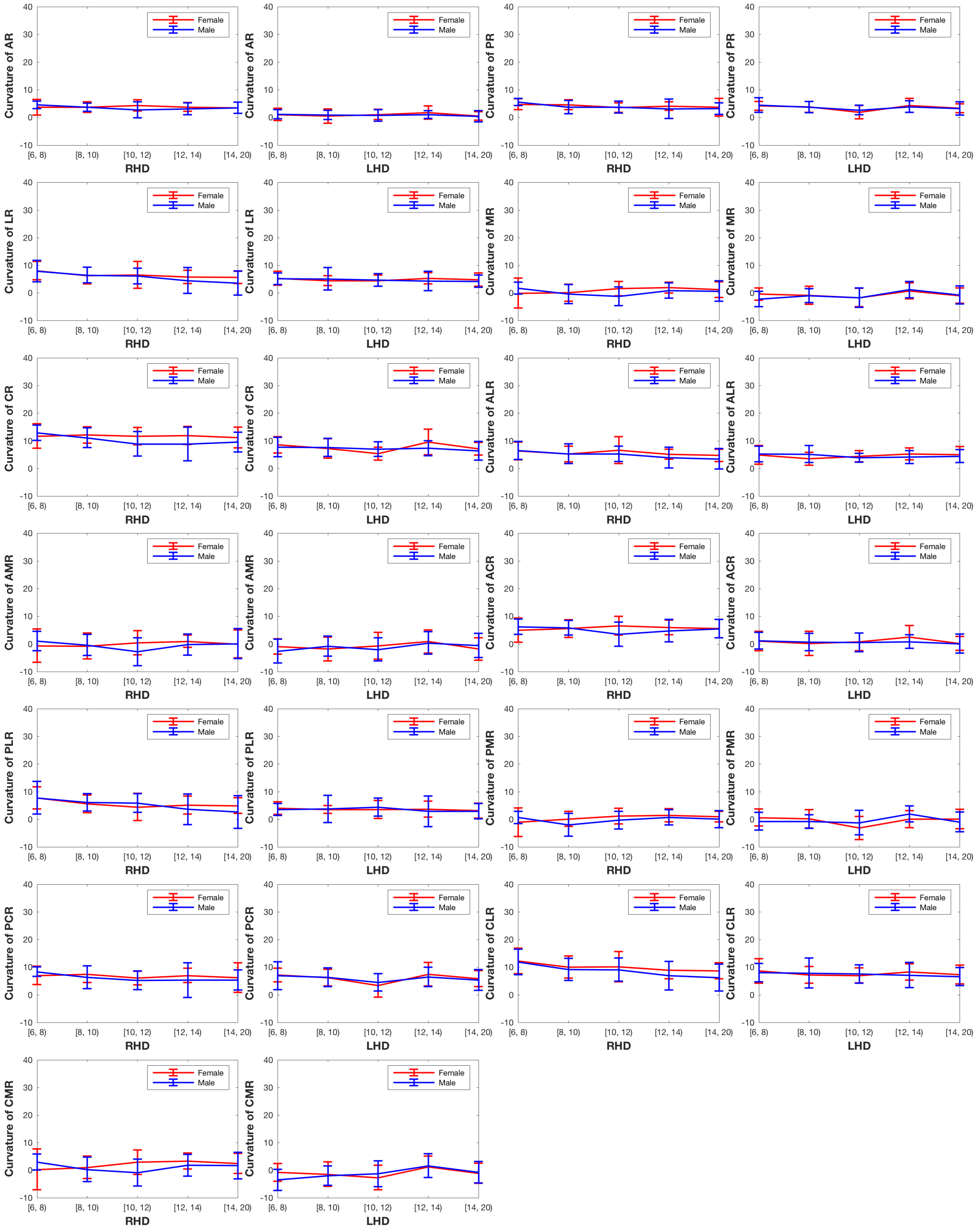

e-Figure 8. Plots of the change in sagittal curvature from end-expiration to end-inspiration in each region for the different age groups and right hemi-diaphragm (RHD) and left hemi-diaphragm (LHD) for both genders. Mean ± one standard deviation limits are also shown.

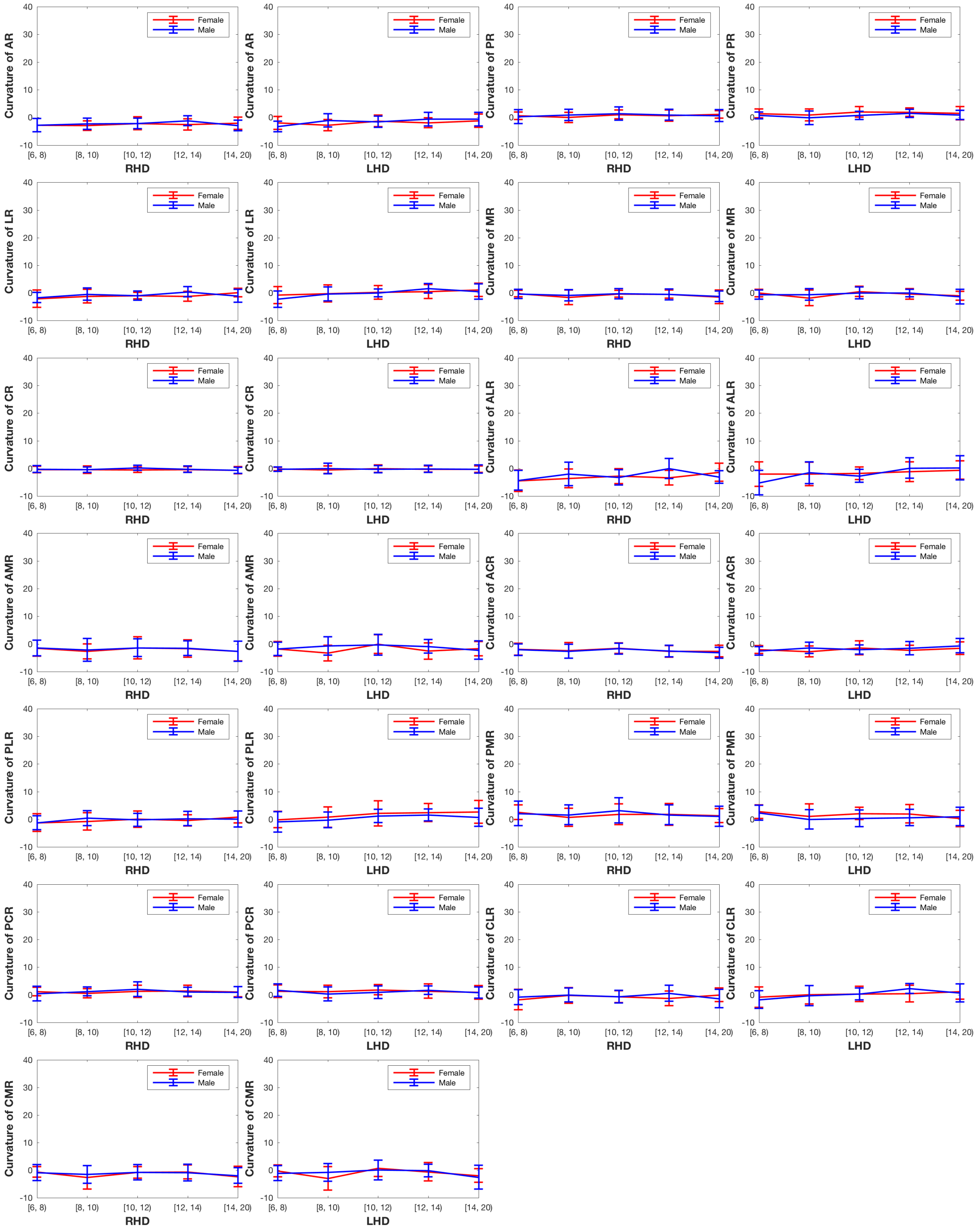

e-Figure 9. Plots of the change in coronal curvature from end-expiration to end-inspiration in each region for the different age groups and right hemi-diaphragm (RHD) and left hemi-diaphragm (LHD) for both genders. Mean ± one standard deviation limits are also shown.

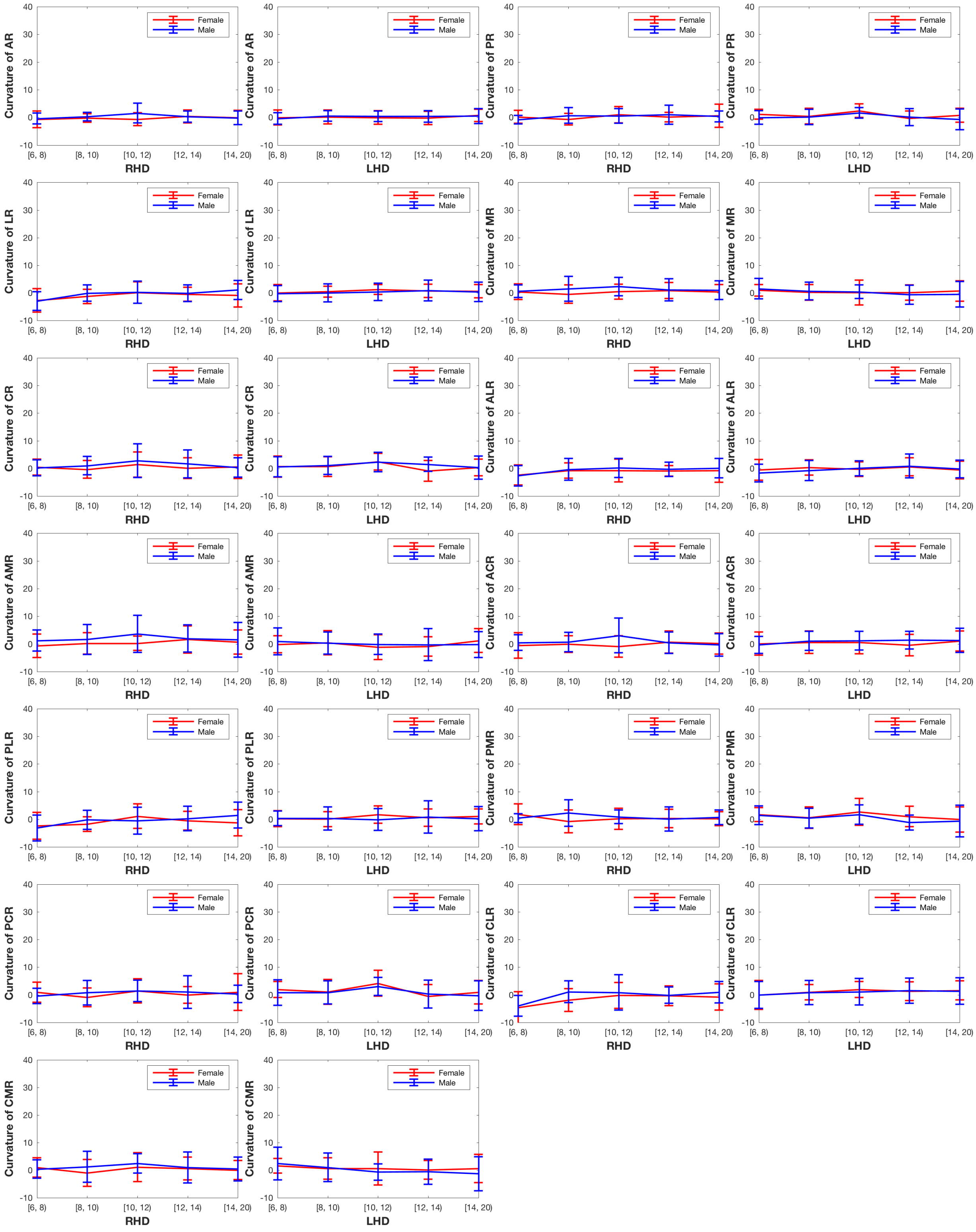
